# The DIRECT study: A roadmap for ctDNA-based risk prediction, molecular profiling and MRD detection in Diffuse Large B Cell Lymphoma

**DOI:** 10.1101/2025.04.14.25325806

**Authors:** Joanna A Krupka, Ilias Moutsopoulos, Natasha H Cutmore, Christopher S Trethewey, Alimu Dayimu, Rebecca Goodhew, Livia Raso-Barnett, Heok Cheow, Lee Elzubeir, Julie Smith, Anver Kamil, Ramona-Rita Barbara, Sally Barrington, Jane Price, Kay Elston, Aleksandra Kolodziejczyk, Silvia Tarantino, Fabiana Mariscotti, Philip Barry, Steven Frost, Nikolaos Demiris, Martin G Thomas, Duane Hassane, Veerendra Munugalavadla, Sateesh Kumar Nagumantry, Mamatha J Karanth, Matthew Ahearne, Nimish Shah, Christopher P Fox, Shubha Anand, Daniel J Hodson

**Affiliations:** Cambridge Stem Cell Institute, University of Cambridge, Cambridge, UK; Department of Haematology, University of Cambridge, Cambridge, UK; Cancer Molecular Diagnostics Laboratory, Department of Oncology, University of Cambridge, UK; Cambridge Clinical Trials Unit – Cancer Theme, University of Cambridge, Cambridge, UK; Department of Statistics, Athens University of Economics and Business, Athens, Greece; Department of Haematology, Cambridge University Hospitals Foundation Trust, Cambridge, UK; Haemato-Oncology Diagnostics Service, Cambridge University Hospitals NHS Trust, Cambridge, UK; Nuclear Medicine, Cambridge University Hospitals NHS Trust, Cambridge, UK; Norfolk and Norwich University Hospitals NHS Trust, Norwich, UK; NoSngham University Hospitals NHS Trust, NoSngham, UK; University Hospitals of Leicester NHS Trust, Leicester, UK; King’s College London, London, UK; Cambridge Blood and Stem Cell Biobank, University of Cambridge, UK; Haematology R&D, AstraZeneca, NY, USA; Haematology R&D, AstraZeneca, South San Francisco, CA, USA; North-West Anglia Foundation Trust, Peterborough, UK; West Suffolk Hospital, Bury St Edmunds, UK; University of Leicester, Leicester, UK; School of Medicine, University of NoSngham, NoSngham, UK

## Abstract

DIRECT was a prospective, multisite study assessing the feasibility and utility of circulating tumor DNA (ctDNA) in 188 patients with aggressive B-cell non-Hodgkin lymphoma using a lymphoma-customized assay and open-source pipeline. CtDNA fraction assessed by copy number alterations in pre-treatment plasma identified high-risk patients more effectively than existing clinical risk scores. In 74.5% of cases ctDNA was equivalent or superior to biopsy for genetic profiling. Patients not suitable for ctDNA genotyping had low tumor volumes and could be predicted from simple clinical factors. Finally, a phased variant-supported minimal residual disease (MRD) assay was predictive of outcomes in all groups analyzed. Patients achieving ctDNA clearance at end of treatment had an extremely low 2-year progression rate (<5%). However, false positive MRD results were common in patients with transformed indolent lymphoma. This study highlights the considerable potential, but also the caveats and limitations, of ctDNA technology when applied to aggressive B-cell lymphoma.

## Introduction

Diffuse Large B Cell Lymphoma (DLBCL) is the commonest form of aggressive non-Hodgkin lymphoma. Although potentially curable with immunochemotherapy, 30-40% of patients will die from their lymphoma. Despite considerable advances in our understanding of DLBCL genomics and extensive clinical trial activity, first-line therapy has improved only incrementally in the last two decades. Circulating tumor DNA (ctDNA) has emerged as an exciting technology with the promise to improve the management of DLBCL by addressing three key challenges that have hampered clinical progress.

First, ctDNA others a molecular assay for risk stratification, allowing high-risk cases to be prioritized for experimental therapy. Pretreatment ctDNA concentration provides a quantitative measure of tumor burden, with a threshold of 2.5 log10 hGE/ml reported to identify high-risk patients^1,2^. However, this threshold was defined using an assay and analytical pipeline that is not widely available. It is notable that a different study, using a different assay, suggested an optimal threshold more than 10-fold higher^3^. This highlights a need to standardize and validate ctDNA thresholds across assays, and to inform how ctDNA burden might be used in conjunction with established clinical risk factors.

Second, DLBCL has emerged as a collection of genetically defined subtypes, each reliant on different oncogenic pathways^4–7^. This underscores the potential for precision medicine approaches in DLBCL^8,9^. However, practical challenges of real-time genetic profiling, including limiting amounts of biopsy material, have hindered the development of trials that tailor treatment to genetic subtype. Previous studies have described the use of ctDNA for plasma-based genotyping of DLBCL^3,10–13^. However, the number of cases and number of variants for which meaningful comparison Has been made between plasma and biopsy genotyping remains relatively small. Key uncertainties remain as to the overall success rates of plasma genotyping, the fidelity of ctDNA-determined mutations, and the factors that predict success or failure of plasma-based genotyping. Furthermore, the use of ctDNA genotyping to enable genetic subtyping with algorithms trained on biopsy tissue has not been systematically evaluated.

Finally, great excitement currently surrounds the use of ctDNA to quantify minimal residual disease (MRD) at the end of treatment (EoT). Ultrasensitive assays such as PhasEd-Seq can quantify ctDNA in DLBCL at concentrations below one part per million (ppm), offering the potential to distinguish those patients who are cured, versus those at high risk of disease recurrence^14^. Ultimately, this might remove the need for lengthy trial follow-up to establish drug efficacy or might enable early intervention with novel therapies in high-risk patients prior to overt clinical relapse. However, the current peer-reviewed literature includes EoT MRD data from only 19 patients, the radiological remission status of whom was not defined^14^. Moreover, PhasEd-Seq is restricted to a single commercial provider and how, or even whether, a similar phased variant-supported MRD assay could be implemented in a routine diagnostic laboratory remains uncertain.

To address these critical questions, we initiated DIRECT, a multicenter, prospective clinical study (NCT04226937) designed to test the feasibility and added value of ctDNA in the clinical management of aggressive B cell lymphoma. We developed a customized, clinical-grade assay and analytical pipeline, focusing our analysis on the three key ctDNA applications: 1) pretreatment risk prediction, 2) plasma-based genetic profiling, and 3) phased variant supported MRD response assessment at the end of treatment. We provide full details of assay development and the end-to-end analytical pipeline with open-source code available for unrestricted use. Our results provide a formal assessment of the value of ctDNA in DLBCL, highlighting the strengths and limitations of current technology, as well as the clinical factors that predict assay performance. The DIRECT study provides a roadmap for the implementation of ctDNA technology in lymphoma in a diagnostic laboratory.

## Results

### Study population and demographics

Between Sept 2020 and April 2023, we enrolled 198 patients with aggressive B cell lymphoma from 6 participating sites within the United Kingdom. Median follow-up for the whole cohort was 25.4 months. A total of 10 patients were excluded from analysis, because of alternative histological diagnosis (n=2), immunochemotherapy not received (n=3), or critical samples missed (n=4) or inadequate (n=1) (**Fig. 1A**). The full analyzable population of 188 patients is referred to as Cohort 1. A more homogeneous subgroup of 165 patients was defined by excluding three patients with Burkitt lymphoma and 20 patients who received non-anthracycline-based chemotherapy. This subgroup of large B cell lymphoma treated with curative intent is referred to as Cohort 2. For each analysis performed, outcomes for Cohort 2 are presented in the corresponding supplementary figure. Full baseline characteristics for each cohort are presented **(Fig. 1B)**. Time-to-tumor-progression (TTP) across international prognostic index (IPI) risk groups followed expected patterns (**Fig. 1C and Fig. S1**).

**Figure 1.**
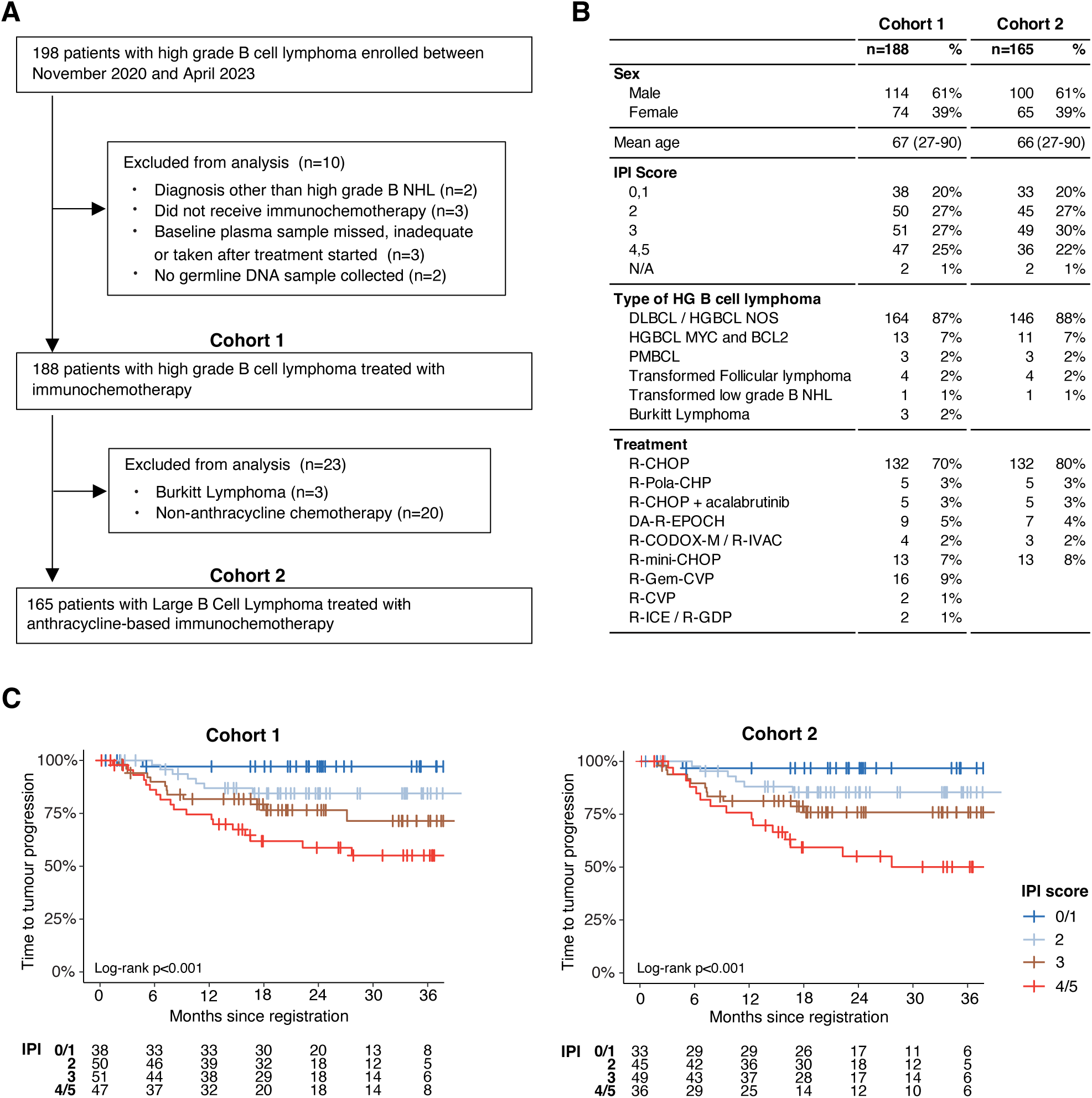
**Patient inclusion and baseline characteristics**. **A**. CONSORT diagram detailing all patients enrolled in DIRECT and their inclusion in the analysis populations Cohort 1 and Cohort 2. **B.** Baseline characteristics of patients within Cohort 1 and Cohort 2. **C**. Kaplan Meier curves showing 6me to progression for Cohort 1 and Cohort 2, stratified by IPI. The statistical tests are based on a two-sided log-rank test. No multiple comparison performed.

### Use of ctDNA for pretreatment risk prediction

We developed a lymphoma-customized, targeted capture-based, next generation sequencing (NGS) panel and analytical pipeline. The latter combined the outputs of four variant callers with a customized noise suppression strategy(**Fig. S2**). Full details are provided in the Methods Section. Panel design, quality control metrics and evidence of the impact of error suppression modules are provided (**Extended Data Table 1&2 and Fig. S3**). In addition to lymphoma driver genes, our panel captured recurrently hypermutated, non-coding regions across the B cell genome. The resulting large number of detected variants (mean 222 per sample) provided a robust measure of variant allele fraction (VAF) across a wide dynamic range (0.99 – 0.001) (**Fig. S4A**). For each sample, ctDNA burden was calculated from the concentration of cell free DNA corrected by the mean VAF, and expressed as log10 transformed, haploid genome equivalents per ml (hGE/ml).

The measurable range of ctDNA burden spanned almost 10,000-fold (**Fig. 2A**) and showed strong correlation with total metabolic tumor volume (TMTV) quantified from baseline PET-CT (R^2^=0.72), and with IPI score (p< 10^-11^) (**Fig. 2B, C**). Consistent with previous reports^1^, patients with pretreatment ctDNA burden above a threshold of 2.5 log10 hGE/ml were more likely to experience progressive disease (PD) at end of treatment (EoT) (**Fig. 2D & Fig. S4B**). This translated into significantly inferior outcomes at 24 months (TTP 68% vs 89% p< 0.001) (**Fig. 2E & Fig. S5&6**). We explored alternative thresholds, observing that clinical risk could be titrated up or down using optimized thresholds of 3.3 or 1.6 log10 hGE/ml respectively (**Fig. S5A, S6A**).

**Figure 2.**
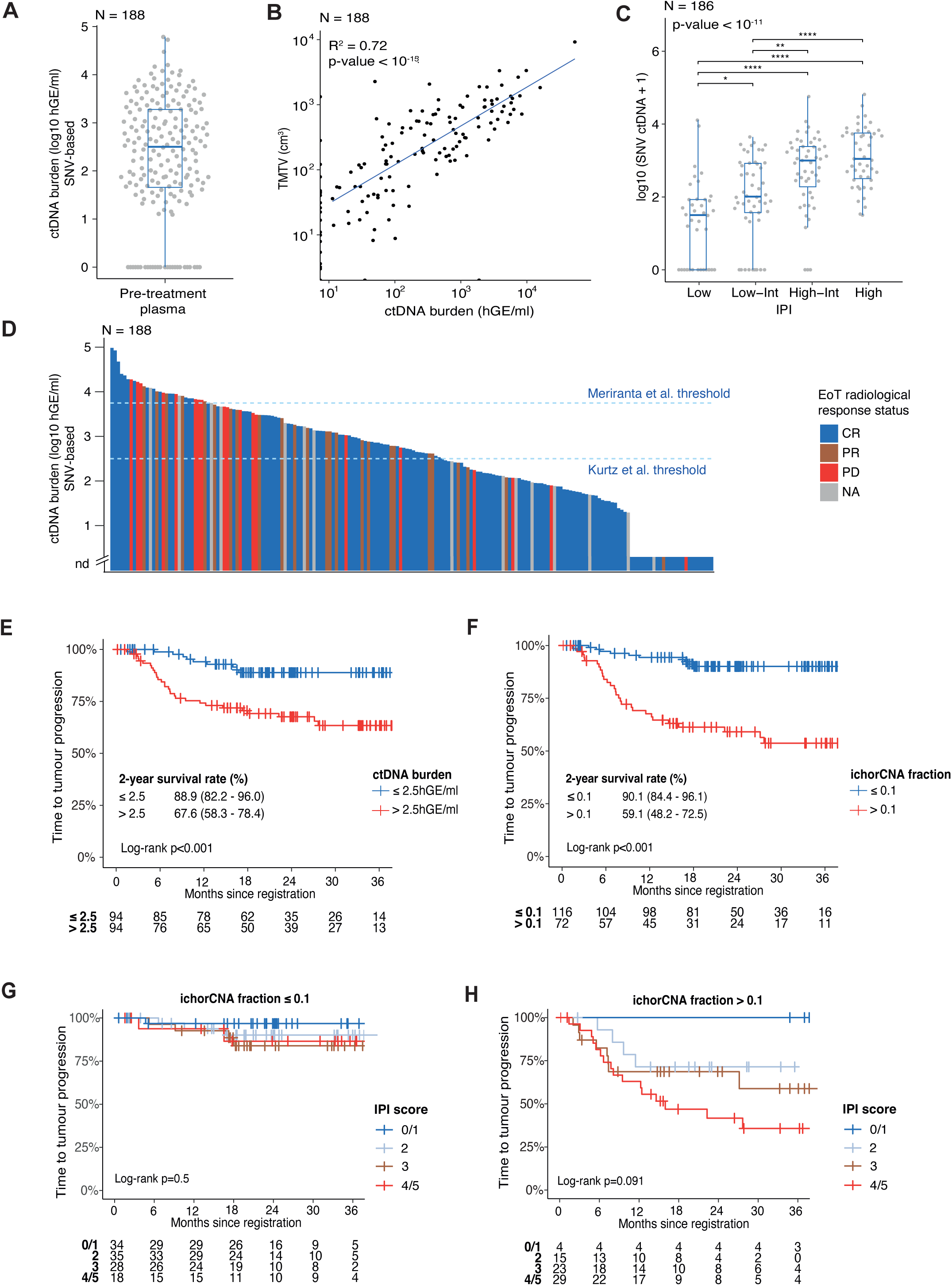
Pretreatment ctDNA as a predictor of clinical outcome. **A**. Distribution of SNV- derived ctDNA burden across all 188 patients in Cohort 1. **B**. Correlation of TMTV from baseline PET-CT with SNV-derived ctDNA burden. R² and p-value, derived from a linear model, are shown. **C.** Association of pretreatment ctDNA burden with IPI risk score (Kruskal-Wallis p- value and significance brackets for post-hoc Dunn test are indicated; * p < 0.05, **p-value < 0.01, ***p-value < 0.001, ****p-value < 0.0001). **D**. Bar chart showing association of pretreatment ctDNA burden with radiological response assessment for all patient in Cohort 1. **E**. Time to progression is shown for all patients in Cohort 1 stratified by pretreatment SNV- derived ctDNA burden using the threshold of 2.5 log10 hGE/ml previously defined by Kurtz et al. **F**. Time to progression is shown for all patients in Cohort 1 stratified by pretreatment ctDNA tumor fraction defined by ichorCNA using a threshold fraction of 0.1. **G-H.** Time to tumor progression is shown for each IPI category amongst patients with low (**G**) and high (**H**) pretreatment ichorCNA-derived ctDNA fraction using a threshold of 0.1. The survival curves are compared with a two-sided log-rank test and no multiple comparison performed.

A concern when applying a fixed threshold across different ctDNA assays is that differences in assay design, sequencing depth, and how low allele fraction variants are handled by differing variant calling pipelines may lead to variation in the calculation of mean VAF^15^. This renders specific ctDNA thresholds applicable only to that individual assay. We therefore tested an alternative strategy to quantify tumor fraction without the requirement to call SNVs. We utilized ichorCNA, a tool originally designed to quantify ctDNA fraction from somatic copy number alteration captured by shallow whole genome sequencing (sWGS)^16^. We discovered that ichorCNA could be applied successfully to targeted panel data (employing both on and oJ-target reads) after normalization to a reference panel of cfDNA derived from healthy individuals (**Fig. S7A**). Shallow WGS and targeted sequencing of the same samples provided highly concordant (R^2^ = 0.94) estimates of tumor fraction (**Fig. S7B**). The ichorCNA-derived tumor fraction and ichorCNA-derived ctDNA burden correlated less strongly with TMTV (R^2^ = 0.18 & 0.39 respectively) than did the SNV-derived ctDNA burden (R^2^ = 0.54) (**Fig. S7C-E**). However, patients with a high ichorCNA-derived tumor fraction (>0.1) experienced particularly poor outcomes at 2 years (TTP 59% vs 90%, p<0.001) (**Fig. 2F, Fig. S5B and Fig. S6B**). Indeed, this approach was at least as predictive of poor outcome as the SNV-derived ctDNA quantification (**Fig. 2E, F and Fig. S7F, G**). IchorCNA is a simple software tool that can be appended to any ctDNA analysis pipeline, regardless of small variant calling strategy, thereby reducing the requirement for cross-assay standardization.

In multivariable analysis, ichorCNA-derived tumor fraction had the highest hazard ratio for tumor progression (TTP HR 14.63) (**Fig. S7H, S7I and Extended Data Table 3**) outperforming IPI and TMTV (TTP HR, 2.07, and 1.17, respectively). Patients with ichorCNA fraction ≤0.1 had excellent clinical outcome regardless of IPI score, whereas those with ichorCNA fraction >0.1 could be further stratified by IPI. (**Fig. 2G, H & Fig. S8**). The combination of ichorCNA-derived ctDNA fraction and IPI provides a simple but powerful metric that could be used in future studies to prioritize the highest risk patients for experimental therapy.

### Feasibility and fidelity of ctDNA for pretreatment genetic profiling

We next assessed the feasibility and fidelity of genetic profiling, comparing pretreatment plasma ctDNA to biopsy tissue. We compared the times taken for samples to reach the central lab since this is a dominant component of turnaround time for a clinical genotyping assay. 98% of pretreatment plasma samples were received by the central laboratory within 7 days (**Fig. 3A**). In striking contrast, no FFPE biopsy material reached the central lab in fewer than 7 days. Furthermore, whereas 99% of pretreatment plasma samples provided adequate ctDNA for NGS library preparation, the FFPE tissue block was either fully exhausted or insufficient for DNA extraction in 26/188 (14%) of cases (**Fig. 3B**). This highlights a hidden, preanalytical failure rate for biopsy-based genotyping and an important logistical advantage to ctDNA-based genotyping when real-time genetic profiling is required.

**Figure 3.**
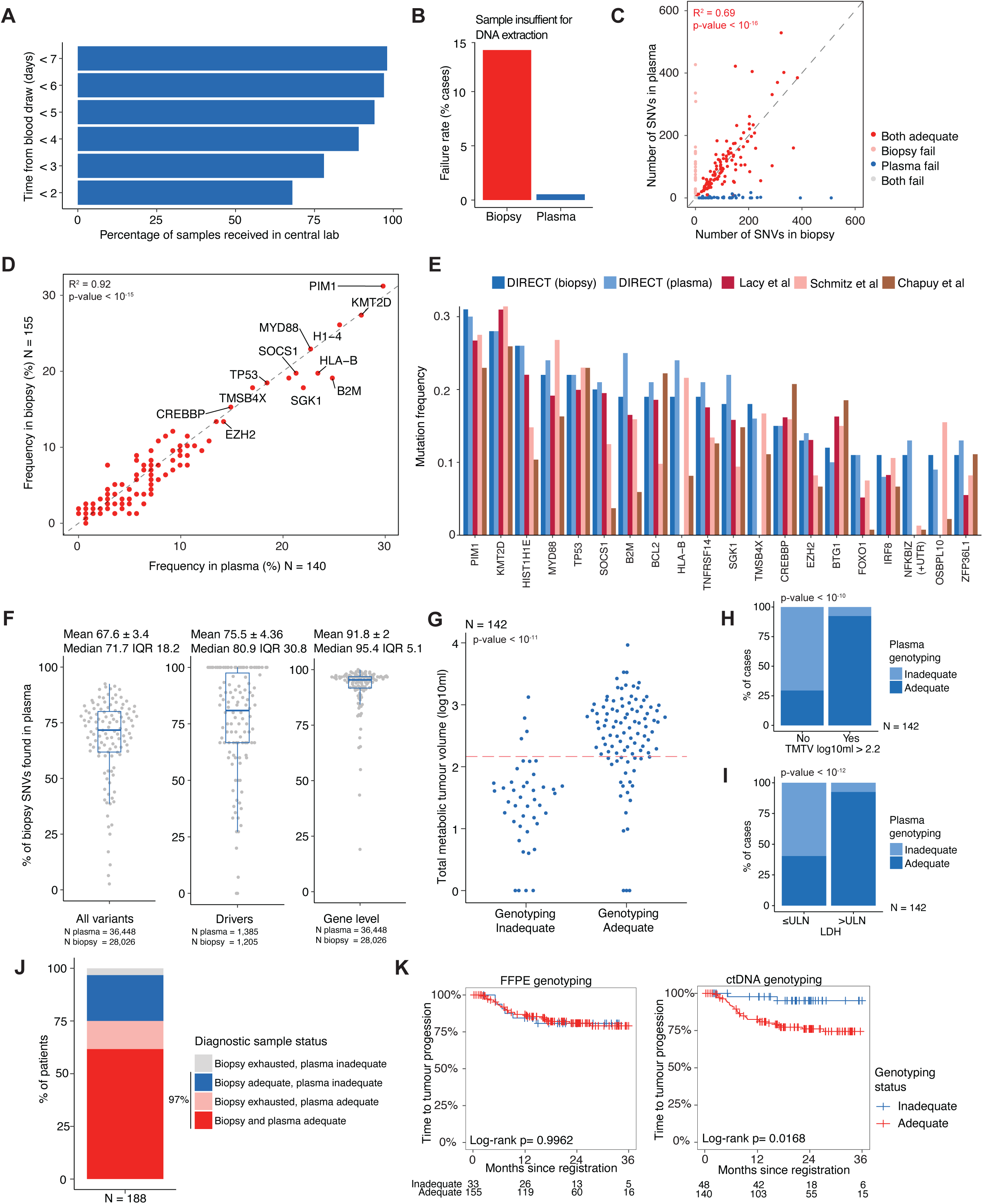
Comparison of pretreatment genotyping using plasma versus biopsy. **A.** Barchart showing the proportion of plasma samples received by the central lab within the indicated number of days from blood draw. **B.** Barchart showing the proportion of samples from either FFPE biopsy of plasma where DNA was insufficient to permit generation of an NGS- sequenceable library. **C.** Total number of non-immunoglobulin variants detected in all 188 patients comparing biopsy and matched pretreatment plasma. patients with failure of either plasma or biopsy genotyping are indicated. R² and p-value, derived from a linear model comparing adequate samples only (red), are indicated in the plot. **D**. Frequency of driver variants identitiied in adequate samples from Cohort 1 comparing plasma and biopsy genotyping. R² and p-value, derived from a linear model, are shown. **E.** Frequency of the most commonly mutated driver genes across DIRECT plasma and biopsy samples compared to published DLBCL sequencing studies. **F.** Percentage of biopsy-proven variants also detected in the plasma, shown for all 141 patients with adequate pretreatment plasma genotyping and a matched FFPE biopsy. Lè panel shows all variants. Central panel shows driver variants. Right panel shows gene level variant calls. The total number of variants considered in each analysis is shown below the plot. **G.** TMTV is shown split by the adequacy of genotyping; n = 142 (cases with missing TMTV values not shown), Wilcoxon p-value. **H, I.** Utitiity of pretreatment clinical factors (TMTV and LDH) to predict success or failure of plasma genotyping; n = 142, Chi-Sq p- values; ULN = Upper Limit of Normal. **J.** Proportion of all patients successfully genotyped using a combination of plasma and/or biopsy-based genotyping. **K.** Association of failed plasma genotyping with improved clinical outcome. The survival curves are compared with a two- sided log-rank test.

To establish the fidelity of plasma-based genotyping, we first compared the total numbers of variants identified in plasma versus the matched biopsy. There was a strong correlation between the number of variants detected in plasma and biopsy (R^2^ = 0.69, average total variants 222 vs 203 respectively **(Fig. 3C)**. The average number of driver mutations was 8.7 in plasma and 8.6 in biopsy (**Fig. S9A, B**). We noted a cluster of plasma samples with very low total variant counts (**Fig. 3C & Fig. S9C**). These samples with fewer than 25 total variants had very few detectable driver variants, little overlap with biopsy genotyping and were deemed plasma fails. These criteria defined a failure rate for ctDNA genotyping of 48/188 (25.6%). For comparison, a failure rate of 33/188 (17.6%) was seen in the FFPE biopsy libraries (considering both sample exhaustion and library failure). 140/188 (74.4%) ctDNA libraries were deemed adequate and used for further analysis. Of these, the distribution of driver genes closely reflected those identified from biopsy (R^2^= 0.92) **(Fig. 3D).** Mutant gene frequencies also mirrored those reported in recent DLBCL sequencing studies **(Fig. 3E)**. All variants identified in either biopsy or plasma are provided as Extended Data (**Extended Data Tables 4 & 5**). To assess how accurately ctDNA recapitulated FFPE tumor genotype, we compared sequencing data from 115 patients with matched and adequate ctDNA and biopsy samples. In total this analysis considered 36,448 total variants and 1,385 driver variants. On average, 71.7% of biopsy-defined variants were also detected in ctDNA. The overlap increased to 80.9% for driver variants and 95.4% for gene-level calls **(Fig. 3F)**.

We further explored the 25.6% of cases where plasma genotyping was inadequate. Forced calling of mutations not identified in plasma suggested that mutant reads were mostly absent, or only present at extremely low counts (**Fig. S9D**). This indicates that a sparsity of mutant DNA molecules captured in the blood draw, rather than lack of sensitivity of the analytical pipeline, was the likely reason for failure of these samples. To identify factors that might predict the success or failure of plasma genotyping, a machine learning approach was employed. This revealed TMTV over 158ml and LDH above upper limit of normal as the two most significant predictors of successful plasma genotyping (**Fig. 3G-I & Fig. S9E-H**); in 95% and 93% cases respectively. We combined these into a simple probabilistic prediction tool that can be used to guide the selection of genotyping strategy in clinical practice (**Extended Data Table 6**).

Overall, we conclude that biopsy and plasma-based genotyping have a failure rate of 16% and 25% respectively. However, they fail for different reasons; sample exhaustion for biopsy, versus low tumor burden for plasma genotyping failure. Plasma provided a suitable source of DNA for tumor genotyping in 74.4% of patients, who could be identified prospectively from simple clinical factors. Overall, 97% of patients could be successfully genotyped using either biopsy or plasma **(Fig. 3J and S10)**. Interestingly, and consistent with low tumor volume as the dominant reason for genotyping failure, patients in whom plasma genotyping failed experienced excellent clinical outcomes **(Fig. 3K)**.

### Use of ctDNA for genetic subtyping of DLBCL

We then compared the ability of ctDNA-derived variants to recapitulate the result of biopsy-driven genetic subtyping using the LymphGen algorithm. This is an important question since LymphGen was trained exclusively on biopsy-derived variants^6^. Biopsy-derived genotyping classified 74% of cases into one of 5 definitive categories, assigning 3% to a composite subtype and leaving 23% unclassified (Other). When genotyped from ctDNA, 67% cases were classified into a definitive subtype, 3% to composite and 30% to Other **(Fig. 4A, Extended Data Table 7).** Cases moving into Other were almost always associated with low number of plasma-derived variants, whereas cases moving into a composite classification were associated with an increased number of plasma variants (**Fig. 4B**). Only a single case changed between definitive classifications (Biopsy-EZB, Plasma-ST2). The veracity of the LymphGen assignment was supported by the results of structural variant (SV) calling. *BCL2*, *BCL6* and *MYC* SVs were called from our NGS assay in 21%, 12% and 7% from biopsy, and 22%, 11% and 11% of cases from plasma (**Fig. 4C**, **Extended Data Table 8)**. Although SV data were not used for our LymphGen classification, almost every *BCL2* SV was detected in a case already assigned to the EZB subtype, and *BCL6* SVs were strongly enriched amongst BN2 cases (**Fig. 4C**). Although SVs were more efficiently identified using biopsy than plasma, the presence of hypermutation (defined as ≥5 variants within 2Kb of the transcriptional start site) faithfully recapitulated the finding of *BCL2* SV (p<10^-15^). This suggests that *BCL2* hypermutation could be used as a robust proxy for *BCL2* rearrangement (**Fig. 4C**). In contrast, *BCL6* hypermutation was seen in both the presence and absence of *BCL6* SV. SVs of *MYC* were too few to establish the predictive value of SHM in this cohort.

**Figure 4.**
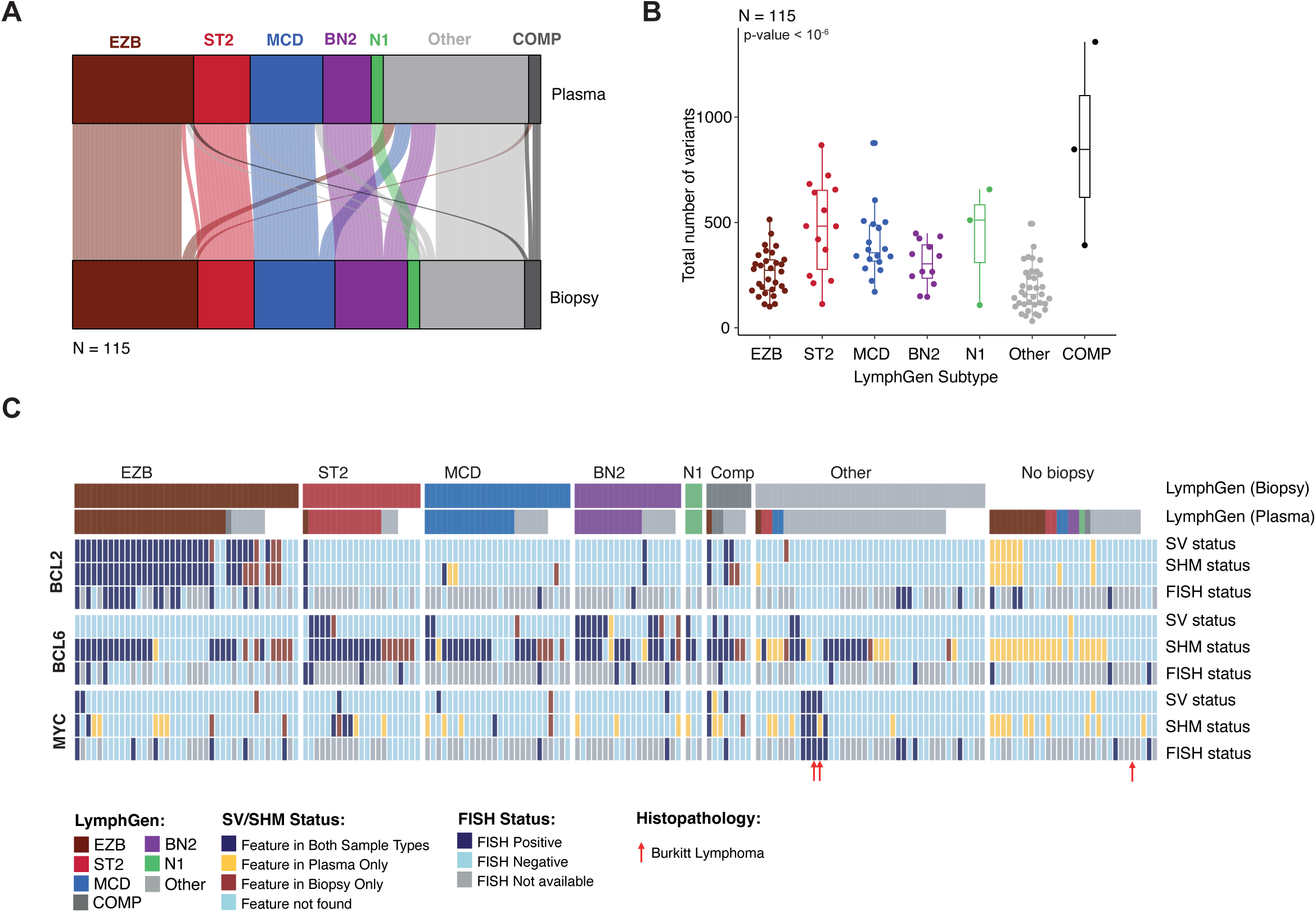
Comparison of genetic subtyping from plasma versus biopsy. **A.** comparison of LymphGen classification from biopsy and plasma-based genotype**. B.** Total number of plasma variants detected in samples assigned to each LymphGen class; Kruskal-Wallis p-value**. C.** Distribution of biopsy and plasma-derived structural variants of *BCL2*, *BCL6* and *MYC* across LymphGen genetic subtypes, including the association with somatic hypermutation status of each gene.

Overall, these data suggest that ctDNA-based genetic profiling permits adequate genetic subclassification using LymphGen, albeit with a slightly higher number of unclassified cases. This makes ctDNA is an attractive strategy to overcome logistic challenges of real-time, biopsy-based genotyping that may enable subtype-directed precision medicine trials in DLBCL.

### Use of a phased variant supported MRD assay for response assessment at EoT

Recent reports suggest that an effective MRD assay in DLBCL must detect tumor DNA with a limit of detection (LoD) approaching one part per million (ppm)^14^. Importantly, the cfDNA extractable from a single 10ml tube of plasma may be limited to just 10^4^ to 10^5^ cell equivalents, mandating the ability to resolve fractions of a single genome equivalent. This can be achieved by tracking many genetic features per cell. However, a LoD of 1ppm falls below the background noise of standard sequencing assays^17^. Tracking variants found in pairs or triplets within individual reads is an elegant strategy to reduce background noise. This phased variant (PV) supported approach is the basis of the recently published PhasEd-Seq assay^14^. We employed the same underlying principle to build a PV-supported MRD assay. Our targeted sequencing panel covered regions of the genome that are densely and reproducibly hypermutated by AID in germinal center B cells. This allowed us to identify many hundreds of PVs from the baseline samples of each patient **(Fig. 5A and S11A-C)**. Indeed, our panel was predicted to capture 90% of PVs that occurred in greater than 5% of patients (**Fig. S11D**). PVs were associated with strong enrichment for mutational signatures of AID activity (**Fig. S11E**) and were identified across all genetic subtypes of DLBCL (**Fig. S11F**). These tumor-specific PVs served as high-confidence, trackable markers at later timepoints.

**Figure 5.**
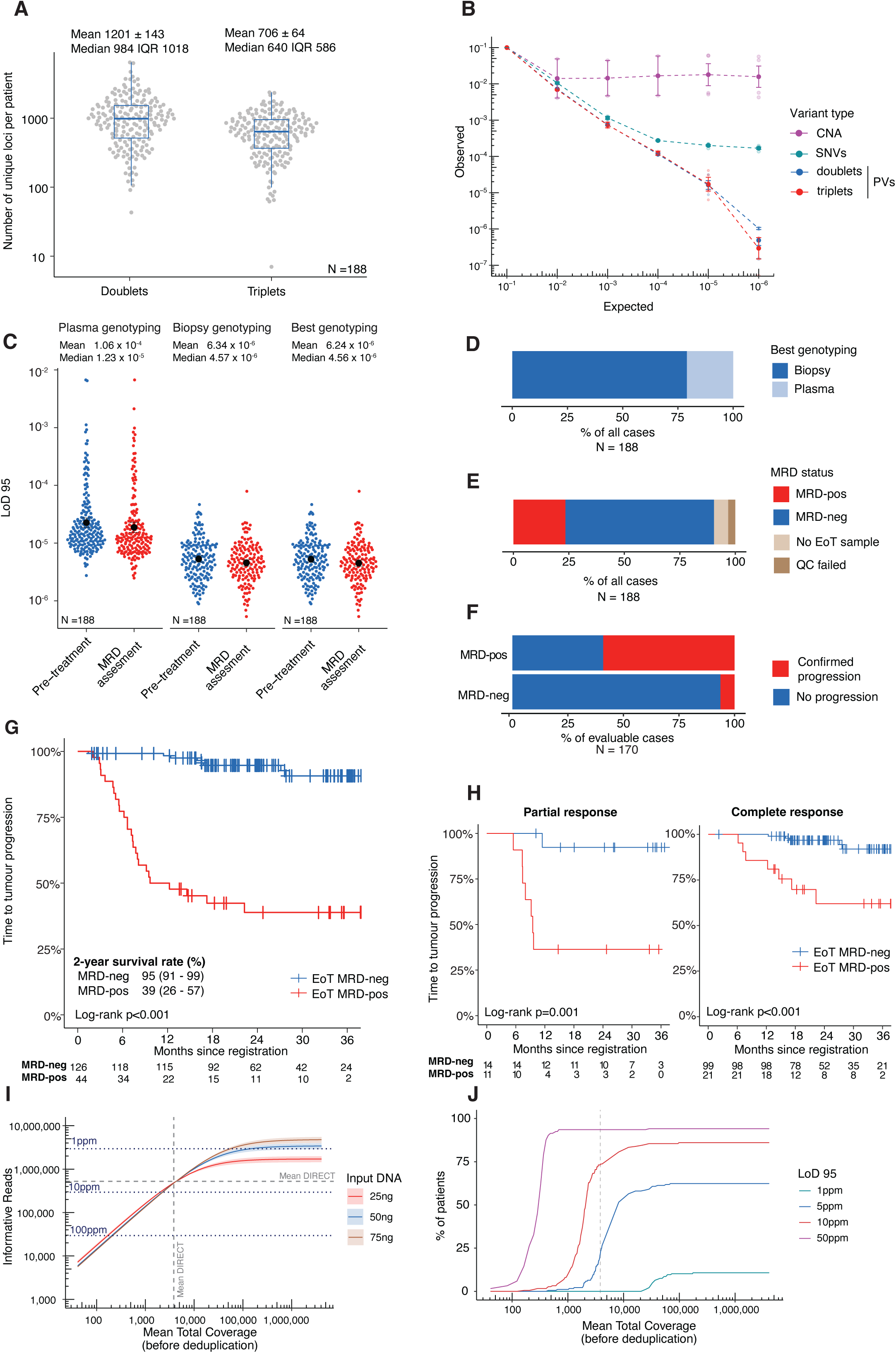
Use of an ultrasensitive PV-supported MRD assay to define EoT response status. **A.** Number of phased variant containing loci (doublets and triplets) identified for each patient at baseline. **B.** Serial dilution of two baseline DIRECT plasma samples into cfDNA from normal plasma, with tumor fraction quantified by ichorCNA, SNV or phased variant (PV) based approaches. **C.** LoD95 is shown for each sample analyzed in DIRECT, comparing a biopsy-based, plasma-based, or best-sample-based strategy to identify trackable phased variants at baseline. **D.** Proportion of all patients for which best genotyping strategy utilized biopsy or pretreatment plasma to identify trackable PVs. **E.** Proportion of all 188 cases for which MRD was positive or negative at EoT. Also shown are the cases that failed QC (informative reads below threshold of 10^5^), or where no EoT sample was available. **F.** Proportions of MRD-pos and MRD-neg cases in which tumor progression was ever seen during study follow-up period. **G.** Time to progression is shown for Cohort 1 patients stratified by MRD status at EoT. **H.** Time to progression is shown for Cohort 1 patients who achieved an EoT radiological response of Partial Response (lè panel) or Complete Response (right panel), stratified by EoT MRD status. The survival curves are compared with a two-sided log-rank test. **I.** Modelling of the predicted LoD95 achieved with the indicated cfDNA input (25ng or 50ng) using the DIRECT panel and cohort at increasing sequencing depths. Dashed vertical line shows mean sequencing depth used in DIRECT. Horizontal dark blue doged lines correspond to a mean LoD95 of 1, 10 and 100 ppm. Grey horizontal dashed line shows the mean LoD95 achieved in DIRECT. **J.** Predicted proportion of Cohort 1 cases who would exceed the indicated LoD95 thresholds as sequencing depth increases using the DIRECT panel and cfDNA input.

Serial dilution experiments confirmed the superiority of a PV-supported approach to quantify extremely low tumor fractions. Whereas SNV based tumor fraction plateaued around 10^-4^, PV-supported quantification remained linear to almost 10^-6^ (**Fig. 5B**). The total number of informative reads (deduplicated reads spanning a trackable doublet or triplet PV) allowed us to derive an individual LoD with 95% confidence (LoD95) for every plasma sample (**Extended Data Table 9)**. The mean LoD95 was strongly affected by the initial genotyping strategy used to define the trackable PVs (**Fig. 5C**). In 80% of cases LoD95 of the EOT plasma samples was superior when using biopsy-informed genotyping (**Fig. 5D**). Using the best genotyping strategy based upon the optimal DNA source provided a median LoD95 across all EoT plasma samples of 4.56x10^-6^ (**Fig. 5C**).

MRD status at EoT strongly predicted clinical outcome (**Fig. 5E, F, G**). Amongst patients with undetectable EoT MRD, 95% remained tumor free after 2 years. In striking contrast, of those with detectable MRD, only 39% remained tumor free after 2 years (**Fig. 5G and Fig. S12**). Importantly, this analysis included patients with a mix of radiological responses and in some cases progression would already have been apparent. Therefore, to establish the added value of the ctDNA MRD status, we restricted our analysis to patients achieving radiological complete response (CR), and separately those with partial response (PR), at end of treatment. This represents two distinct scenarios where MRD may add new information. In both scenarios, patients with MRD negativity had an extremely low rate of recurrence (3% and 8% respectively). In contrast, MRD positivity was associated with disease recurrence in 38% and 64% of patients in CR and PR respectively (**Fig. 5H & Fig. S12)**. Examining the characteristics of MRD positive patients without relapse, we noted that 3 of 4 (75%) cases of transformed follicular lymphoma enrolled in our study remained MRD positive after therapy without relapse. When inspecting individual PVs, these showed robustly detectable PVs at EoT (**Fig. S13**). This suggests detection of a persisting indolent lymphoma subclone that may be independent of the transformed lymphoma. Among the remaining cases with detectable EoT MRD but no relapse, we noted that 2 received subsequent consolidation radiotherapy, and 4 had a follow-up period of less than 2 years.

We also investigated the small number of patients who relapsed despite negative MRD. Resequencing of these 5 patients at greater depth did not convert these patients to MRD positive status. Therefore, we modeled the factors that limited sensitivity of our assay and what would be needed to achieve a LoD95 of 1ppm across the majority of our cohort. Since our panel already captured the vast majority of recurrent PVs in the DLBCL genome (**Fig. S11A-D**) we focused on sequencing depth and cfDNA input. This analysis revealed that in most cases, deeper sequencing yielded only trivial improvements in LoD95, with most libraries already approaching sequencing saturation (**Fig. 5I, J**). Instead LoD95 was limited in most cases by input DNA (at the EoT, 15% of samples yielded less than 25 ng of DNA available from a 10 mL blood tube). Our model showed that increasing LoD95 to 1ppm would require 2-3-fold increased DNA input combined with 20-40-fold increased sequencing depth.

Overall, these findings confirm the utility of PV-supported MRD as a response assessment strategy. This approach adds valuable information beyond current radiological assessment, with enhanced sensitivity to detect residual disease in patients achieving CR and offering an orthogonal approach to resolve ambiguous imaging in patients not in CR. Our data show how the limit of detection of this assay is unique to each patient, heavily impacted by the initial genotyping strategy and LoD95 was ultimately limited by the amount of cfDNA captured in the blood draw. The assay has a high negative predictive value, but transformed indolent lymphomas represent a specific scenario where caution should be exercised when interpreting positive MRD status.

## Discussion

A series of recent studies have engendered considerable excitement over the potential for ctDNA to transform the management of DLBCL^1–3,10–14,18,19^. Specific applications include baseline risk prediction, tumor genotyping, and ultrasensitive MRD quantification. However, our current understanding of ctDNA utility in the clinical management of lymphoma remains in its infancy. Published studies have been limited by small cohort size and the predominant use of retrospective, archival material. Importantly, available data have been generated using assays and analytical pipelines whose availability is often restricted to a single laboratory or single commercial provider.

Data from the DIRECT study confirm previous reports that pretreatment ctDNA burden can be used to identify high-risk cases, conferring additional prognostic information over existing clinical or radiological risk factors. However, selecting a standardized ctDNA threshold that can be applied across different assays is more complex. Absolute quantification of ctDNA requires the total cfDNA concentration to be adjusted by the average VAF, a metric influenced by the sensitivity of individual assays and how variant calling pipelines handle low allele frequency, subclonal variants. A ctDNA threshold of 2.5 log10 hGE/ml was robustly validated for the CAPP-Seq assay^1,2,20^. However, this is assay is not widely available, requiring individual labs to develop their own assay and analytical pipeline, in turn requiring them to validate their own ctDNA threshold. Indeed, a threshold more than 10-fold higher was defined by a second group, using a different assay^3^. We investigated an alternative approach using ichorCNA^16^, a simple software tool applied in parallel to, but not dependent upon SNV calling to quantify ctDNA fraction. Although developed for use with sWGS, we show that when normalized to a panel of normal plasmas, it can be applied to targeted panel sequencing. Importantly, much of the data used by ichorCNA comes from oJ-target reads, reducing the impact of alternative panel designs on the quantification of ctDNA fraction. Moreover, we find that ichorCNA-inferred high tumor fraction is better able to identify high risk patients than the standard SNV-inferred ctDNA burden. We hypothesize that the dependence on copy number disruption may capture an additional aspect of high-risk tumor biology, beyond simply tumor burden. Indeed, ichorCNA-derived tumor fraction above 0.1 was the pretreatment variable most predictive of poor risk in multivariable analysis. This metric provides prognostic information over and above standard IPI factors and, when combined with IPI, it provides a robust and powerful strategy to identify patients for whom standard therapy may be inadequate. Such an approach will enable future clinical trials where treatment is stratified by baseline clinical risk.

Previous studies have also suggested that ctDNA may be used for baseline genetic profiling of DLBCL^10–13,20^. However, biopsy-plasma comparisons have only been made on comparatively small number of patients and assessed only a small number of variants. Moreover, the extent to which a plasma genotyping approach would be successful, or the factors that might predict success have not been formally studied in a prospective clinical study. The DIRECT study reveals that biopsy and plasma genotyping had a non-trivial failure rate (16% and 25% respectively). However, failure was due to different reasons. Biopsy genotyping commonly failed due to exhaustion of the tissue block leaving insufficient material for DNA extraction. Given the dominant diagnostic approach increasingly employs core needle biopsy, the competition for limiting quantities of biopsy material becomes an even more pressing problem. In contrast, plasma genotyping failed in patients with very low tumor volume. This reflects the tiny number of such scarcely abundant mutant DNA molecules captured within the 10ml blood draw. Our findings suggest that successful genotyping of low tumor volume patients will require greater DNA input than can be captured in a 10ml blood draw. We show that the probability of plasma genotyping success can be predicted from simple clinical factors – in particular, TMTV and LDH, and provide a simple tool that can be used to guide selection of the most appropriate genotyping strategy (**Extended data table 6**). For most patients however, we find that plasma provides an appropriate source of tumor DNA for genotyping that faithfully reflects the results of biopsy genotyping and allows application of genetic subtyping algorithms such as LymphGen. Notably, we only saw a single instance when plasma and tumor profiling led to different, high-confidence subtype assignment. Finally, we highlight considerable logistical advantages to plasma genotyping especially where histological and molecular analyses are often performed in separate laboratories, creating competition for timely access to the tissue block. Indeed, plasma-based genotyping may be the most practical strategy to enable a clinical trial where targeted therapy is directed by pretreatment genotype.

The ability to distinguish patients who are cured at the end of treatment from those at risk of relapse could enable progress in three ways: ctDNA was recently approved as a surrogate trial endpoint in myeloma, potentially removing the need for protracted follow-up and unnecessary imaging-related radiation exposure. Detection of persisting MRD might also allow earlier intervention with CAR-T or bispecific antibody therapies that may have greater efficacy and less toxicity in the absence of overt relapse. Finally, de-escalation of immunochemotherapy for patients who achieve MRD negativity early into treatment others the potential to reduce toxicity burden for those with an excellent prognosis. Use of MRD for these decisions requires an assay able to detect extremely low levels of residual tumor. Current NCCN guidelines recommend the use of a ctDNA assay with sensitivity of one part per million to assess MRD at EoT. Such sensitivity has only been reported by the PhasEd-Seq assay, which was also reported to be 100% predictive of cure or relapse in MRD negative and positive patients^14^. However, the number of patients in that retrospective study was small and their radiological remission status was not provided. Moreover, PhasEd-Seq assay can only be accessed through a single commercial provider and the complex analytical pipeline required to run an equivalent assay in-house is not openly available. Therefore, the practicalities of achieving this degree of MRD sensitivity have not been comprehensively studied. We developed an independent phased variant supported MRD assay and assessed its performance in a prospective clinical study. Full details of assay design and open-source, end-to-end analytical code are freely available under open-source license. We highlight how the limit of detection of this assay is patient specific, with median LoD95 of 4.56x10^-6^. Patient-to- patient differences in LoD95 were determined in large part by the number of trackable PVs identified at baseline. This explains the superiority of biopsy-informed versus plasma-informed approaches for identification of trackable PVs, enhancing average LoD95 almost 10-fold. Modelling of WGS data showed that our panel captured 90% of PVs occurring in 5% or more of biopsies, suggesting that improvement to the design of a universal panel is unlikely to improve LoD95 any further. The design of patient-specific, WGS-informed MRD panels would be one approach to increase the number of identified PVs but would have clear logistical disadvantages. We sequenced our MRD libraries to a median deduplicated coverage of 2,500x. Deeper sequencing led to marginal sensitivity gains due to saturation of libraries constructed from low (25ng) input cfDNA. We used 25ng cfDNA as this could be extracted from a 10ml blood draw in at least 85% of patients at EoT. Our subsequent modelling predicted that increasing DNA input at least 2-4-fold, combined with increased sequencing depth of at least 20-40-fold would be required to achieve LoD95 of 1ppm across the whole cohort.

However, despite only achieving median LoD95 of 4.56x10^-6^ in our cohort, MRD negativity was associated an extremely low rate of relapse (2-year progression only 5%). This low false negative rate, challenges the necessity to target an LoD95 of 1 ppm, especially given the vast increase in sequencing cost required to achieve this level of assay sensitivity. MRD negativity was equally predictive in patients with radiological CR as well as those with non-CR, providing a robust and orthogonal metric to interpret ambiguous radiological data, potentially avoiding potential harms of unnecessary biopsy or radiation exposure. Importantly, not every patient with persisting MRD positivity at EoT relapsed during the study period. Amongst patients achieving CR at EoT only 46% relapsed. This may reflect the limited follow-up window. However, it is intriguing that three of four cases of transformed lymphoma had positive MRD at EoT without subsequent relapse. This suggests the persistence of an underlying indolent lymphoma clone and acts as a caution when considering pre-emptive intervention in an MRD-positive patient known to have transformed from indolent lymphoma.

In summary, we report the results of a prospective clinical study specifically designed to assess the feasibility and utility of ctDNA in the management of DLBCL using an open-source assay and pipeline that could be deployed in any diagnostic laboratory with NGS capability. Our data support and build upon existing literature and confirm the added value of ctDNA beyond established clinical and radiological metrics. Importantly, they reinforce the power of a phased variant supported ctDNA assay at EoT to define a patient group with an extremely low rate of recurrence. Data from DIRECT highlight limitations of ctDNA technology in DLBCL and caveats to data interpretation, relevant both for clinical trial design and implementation in routine practice.

## Methods

### Patient recruitment and study eligibility criteria

Patients were enrolled into the DIRECT study (NCT04226937). This study was sponsored by Cambridge University Hospitals NHS Foundation Trust and the University of Cambridge. Inclusion criteria were as follows:

- Have given written informed consent to participate.
- Age ≥ 18 years at the time of consent.
- Histologically confirmed diagnosis of previously untreated high-grade B cell lymphoma.
- Planned to receive immunochemotherapy as first-line therapy, e.g. R-CHOP therapy.
- Planned or completed standard of care imaging (CT or PET-CT) before commencing treatment.
- Able to give blood. Exclusion Criteria were as follows:

The presence of any of the following will preclude participant inclusion:

- Unable to receive immunochemotherapy as first-line therapy due to co-morbidity or personal choice.
- Patients who have already started high dose steroids as a treatment for their lymphoma.
- Known diagnosis of infectious blood-borne virus e.g. Hep B, Hep C or HIV.

The DIRECT study was reviewed and given favorable opinion by East of England – Cambridge Central Research Ethics Committee (20/EE/0068) and the UK Health Research Authority. All patients provided written informed consent prior to participating. The study was performed in accordance with the spirit and the letter of the declaration of Helsinki, the conditions and principles of ICH E6 Good Clinical Practice, the protocol and applicable local regulatory requirements and laws.

### Radiological response assessments

Results of all standard-of-care PET-CT and CT radiology assessments were collected and entered by sites into the electronic trial database. We used a series of rules to define an integrated end of treatment (iEoT) response as follows:

1. Any scan up to and including EoT scan reported as Progressive Disease. Status = PD
2. Clinically reported disease progression prior to EoT Scan. Status = PD
3. EoT PET reported as D1, D2, D3 or as Complete Response. Status = CR
4. Interim PET scan reported as D1-3 or Complete Response, with no subsequent clinical or radiological report of Progressive Disease. Status = CR
5. EoT CT is reported as Complete Response. Status = CR
6. No EoT scan performed within window, but “late EoT scan” performed within 6 months following completion of treatment reported as D1-3 or Complete response. Status = CR
7. EoT PET reported as D4. Status = PR
8. EoT PET reported as D5, but site have not reported as Progressive Disease. Status = PR
9. EoT PET reported as Partial Response without Deauville score. Status = PR
10. Interim PET reported as D4/D5 or Partial Response with no subsequent EoT scan and site have not reported as Progressive Disease. Status = PR

### Measurement of TMTV

Total metabolic tumor volume was measured using the international benchmark method, including tumor with SUV≥4 and minimum volume of 3mls using standard software for clinical reporting^21^. Measurements were made by radiologists at the participating PET centers where the scans had been acquired using the standard of care acquisition and reconstructions.

### Definition of survival metrics

Time to tumor progression (TTP) was calculated from the date of participant registration to the confirmation of tumor progression or relapse that was confirmed by either radiology or biopsy whichever occurs first. Deaths in the absence of confirmed tumor progression or relapse were censored. Progression Free Survival (PFS) was calculated from date of participant registration to date of the first disease progression or date of death from all causes, whichever occurred first. Patients who were alive and clinically well (no relapse or progression), were censored at the date of last known alive. Overall Survival (OS) was calculated from date of participant registration to date of death from all causes. Patients were censored for this endpoint on the date last known alive.

### Definition of study timepoints

The EoT ctDNA sample was defined as the sample taken at the first visit after last cycle of induction chemotherapy. For a small number of patients where an EoT sample was not collected, we used the C3D1 sample if the patient has already achieved MRD negative status.

### Processing of blood and plasma samples

Plasma was obtained from whole blood drawn into PAXgene Blood ccfDNA Tubes (768115 / QIAgen) and shipped to the central laboratory within 7 days. PAXgene tubes were centrifuged at 1900g for 15 minutes to separate the plasma phase, prior to a second centrifugation of the isolated plasma at 1900g for 10 minutes to remove residual cellular debris. Plasma was stored in FluidX tubes as 6mL aliquots at -70°C. Granulocytes (used for germline DNA isolation) were purified from 1mL of whole blood drawn in K3EDTA (Monovette) blood tubes using EasySep Direct Human Pan-Granulocyte Isolation Kit (STEMCELL Technologies, 19659) according to manufacturer protocol. Isolated granulocytes were resuspended in 210µL (≈ 1 X 10^7^ cells) PBS prior to storage and subsequent DNA extraction.

### DNA extraction from plasma

Isolation of cfDNA from plasma was performed using the QIAsymphony SP (QIAgen / 9001301) and QIAsymphony® DSP Circulating DNA Kit (QIAgen / 937556) to manufacturers protocol. Plasma in 6mL aliquots were thawed at 30°C for 30 minutes prior loading to the QIAsymphony SP. Following automated DNA extraction, samples were eluted in 60µL of elution buffer and quantified using the Qubit 3 High Sensitivity Assay (ThermoFisher), prior to storage at -70°C. Towards the end of the study, cfDNA was extracted from plasma using the Maxwell (Promega) RSC ccfDNA Plasma LV Kit (large Volume) (Promega AS1840) on the automated Maxwell® RSC 48 (Promega) instrument, to manufacturer’s instructions. Plasma in 6mL aliquots were thawed at 30°C for 30 minutes, prior to addition of binding buffer and 100µL resin before being incubated at room temperature on an orbital mixer (Fisherbrand) for 45 minutes. After incubation, samples were centrifuged 1000g for 2.5 minutes to pellet the DNA coated resin. Following removal of the supernatant, the resin pellet was re-suspended in 500 µL binding buffer and loaded onto the instrument for cfDNA isolation. Upon completion, cfDNA was eluted by the instrument in 60µL NGS Elution Buffer (Promega) and quantified using Qubit-3 High Sensitivity DNA kit (ThermoFisher) to manufacturer’s instructions.

### Extraction of germline DNA from Granulocytes

Genomic DNA was isolated from purified granulocytes with the QIAGEN QIAamp^®^ DNA blood mini kit (Qiagen 51104) according to manufacturer’s instructions. For initial cellular lysis, 20μL of protease was added to 210µL purified granulocytes in PBS, followed by 4 µL RNase A and 200μL of buffer AL prior to incubation at 56°C for 10 minutes. Following QIAgen standard extraction procedure, DNA was eluted in 100 µL of buffer ATE after repeat re-saturation of the column with the eluate to increase yield. DNA was quantified using Qubit 3 DNA Broad Range kit and stored at -70°C.

### DNA extraction from biopsy

For each biopsy, ten slides were cut for diagnostic FFPE tissue blocks for DNA isolation using the QIAgen DNA FFPE kit (Qiagen / 56404) to manufacturers protocol. Prior to scraping of slides for nucleic acid extraction, formalin- fixed, paraffin-embedded tissues underwent local microscopic analysis to mark neoplasia against background healthy tissue to increase the analytical tumor fraction. Slides were deparaffinized using Xylene followed by sequential ethanol washes prior to scraping and macro-dissection. Dried deparaffinized tissues were digested with 20µL of Proteinase K with buffer ATL in a thermomixer at 56°C overnight. Samples were further incubated at 90°C for 1 hour to partially reverse formaldehyde modification of nucleic acids from initial fixing. Following QIAamp DNA FFPE Tissue protocol, DNA was eluted in 50µL EB buffer, quantified using Qubit 3 Fluorometer Broad Range Assay (ThermoFisher) and stored at -70°C.

### Design of targeted panel

We employed a custom-designed 559kbp capture panel from TWIST (Design-ID: TE- 91289462). This captured 130 DLBCL driver genes, translocation sites for *MYC*, *BCL2* and *BCL6* and somatic hypermutation hotspots within immunoglobulin and other hypermutated loci. (**Extended Data Table 1**). The panel was designed by reviewing the published lists of mutations from several large DLBCL sequencing studies^4,5,7^. The translocation sites were guided by published rearrangement co-ordinates within *BCL2*, *BCL6*, *MYC* and immunoglobulin loci^4,22^. Somatic hypermutation hotspots in 5’ regions of the gene loci were taken from previously published whole genome sequencing data^23^.

### Preparation and sequencing of targeted NGS libraries

Libraries were prepared following TWIST library preparation procedure following mechanical fragmentation of genomic DNA samples. Prior to preparation, genomic DNA from FFPE tissue and granulocytes were fragmented using Covaris M220 Ultrasonicator for 200 seconds (Peak power 75, duty factor 10, cycles 200, temp 20°C) to achieve fragment lengths of approximately 200bp in length. Following end-repair and A-tailing, 70 ng of fragmented gDNA or 25 ng of cfDNA was ligated with Integrated DNA Technologies Xgen-UMI adapters prior to hybridization with TWIST custom lymphoma panel (606kbp) at 70°C for 12 hours as plexes of 8 samples. Quality control of library pools was performed with Agilent 4200 TapeStation with D5000 High Sensitivity ScreenTape kit and quantified with Qubit Fluorometer. Samples were sequenced with Illumina NextSeq 2000 (S2 or S3 Kit) and NovaSeq-X 25B kit to achieve a mean depth of 1311 for gDNA samples and 2563 for cfDNA samples. Run performance was verified using Illumina sequencing analysis viewer. FASTQ files were further interrogated using FastQC (Babraham bioinformatics) with multQC to verify run performance.

### Bioinformatics analysis

#### Open-source computational framework

The Automated Ultrasensitive Lymphoma Evaluation (AULE) pipeline was built based on the British Colombia Cancer Research (BCCRC) Lymphoid Cancer Research (LCR) pipeline, developed by the Morin lab (https://github.com/LCR-BCCRC/lcr-modules).

AULE was developed to utilize Snakemake workflow management system and to follow the same modular structure as LCR- BCCRC pipeline. Full code together with user manual was released as an open-source software on GitHub (https://github.com/Hodson-Bioinformatics/Aule).

#### Preprocessing and alignment

##### AULE module: aule_preprocessing

We followed the <u>Genome Analysis Toolkit (GATK) Best Practices</u> workflow for processing raw FASTQ file and generating clean BAM files for variant calling and further analysis. We introduced a few modifications to support the integration of Unique Molecular Identifiers (UMIs), that were provided in a separate FASTQ file. After raw R1, R2 and UMI FASTQ files were converted to unmapped bam files with (Piccard FastqToSam), the reads were annotated with their respective UMIs using fgbio AnnotateBamWithUmis. The reads were aligned to the reference genome (hg38), with BWA-MEM.

#### Quality control

##### LCR-BCCRC modules: qc, picard_qc

Quality control metrics were calculated following best practices for information reporting in genomics experiments (ref: https://doi.org/10.1182/blood.2022016095). Metrics were obtained through the gatk Collect*Metrics, samtools stats and the picard Collect*Metrics commands applied to aligned bam files. Coverage, fragment size, duplication rate, sequence and mapping quality metrics were produced for each sample and were presented by sample type (Germline, FFPE Biopsy, Plasma), by hospital site, and by the date of sequencing (**Fig. S3A**).

#### Unique Molecular Identifiers (UMIs) processing

##### AULE module: aule_preprocessing

We grouped reads originating from the same DNA molecule and assessed the concordance of base calls at corresponding positions. Reads that were not properly paired, or that represented secondary, supplementary, or unmapped alignments, were excluded from the analysis. We validated mate read mapping quality and the mate CIGAR string using fgbio SetMateInformation. Reads sharing the same end position and UMI sequence - indicating that they were derived from the same DNA molecule - were grouped into UMI families with fgbio GroupReadsByUmi using the directed adjacency method. The grouped reads were then collapsed into a single consensus sequence with fgbio CallMolecularConsensusReads, with overlapping read pairs excluded from consensus calling (--consensus-call-overlapping-bases false). All other parameters were maintained at their default values, and UMI families of size 1 were retained. The consensus BAM was then re-aligned to the reference genome (hg38) as described previously, and the resulting aligned BAM was merged with the original consensus BAM using GATK MergeBamAlignment to retain positional consensus calling metrics. Finally, discordant bases - either due to a lack of consensus within the UMI family or the read pair - were converted to N using fgbio’s CallOverlappingConsensusBases with the parameters --agreement-strategy PassThrough and --disagreement-strategy MaskBoth.

#### Somatic Single Nucleotide Variants (SNVs) calling

##### LCR-BCCRC modules: slms3 AULE modules: aule_pon

We performed SNV calling using four different variant callers - Sage, LoFreq, Mutect2, and Strelka2 (SLMS-3 pipeline), as previously described (Thomas et al. 2023; Hilton et al. 2023). All variants were called in matched germline mode. We generated a customized panel of normals (PON) using cell-free DNA from 51 healthy donors. A combined PON VCF file, used to filter out likely non-tumor-specific calls (i.e., germline variants and sequencing artifacts), was created with GATK CreateSomaticPanelOfNormals. All variant calling filters were maintained as per the original workflow except Strelka2 where the minimum SomaticEVS was set to 0.2.

For FFPE biopsies, only high-confidence variants - those called by at least three variant callers (3+ variants), were included. For calling SNVs in ctDNA we developed customized error suppression and introduced a 2-stage variant filtering to increase the variant calling sensitivity for low tumor burden cases.

#### Error suppression and ctDNA SNVs filtering

##### AULE modules: aule_error, aule_plasma, aule_out_tables

Background error rates for each panel position were estimated from the healthy samples. The errors considered were: UMI errors, where a UMI “family” is discordant between its members, R1/R2 error, where the R1 and R2 reads are overlapping and discordant at a location, and directional errors, where the base at a position does not match the reference. The rates for all error types were calculated by a custom python script using pysam: A bam file containing UMI collapsed reads produced by fgbio CallMolecularConsensusReads was used as input, aligned and sorted by query name. For each read pair, the aligned genomic positions were extracted, as well as the per-position consensus depth and consensus errors (cd & ce tags respectively), and the sequence agreement for the positions that the R1 and R2 reads are overlapping. The following metrics were tallied for every position in the panel as the reads in the bam file were parsed:

- Total number of UMI families of size 1 (trivial), 2+ (nontrivial).
- Number of UMI errors (ce tag in the bam file).
- Total number of overlapping R1 & R2 reads.
- Number of discordant R1 & R2 reads (different base in the overlap).
- Number of reads & UMI families containing each base (A, T, C, G).

These were then used to compute the UMI error rate (# UMI errors / # UMI families), R1/R2 error rate (# R1/R2 discordant / # R1/R2 overlapping reads), and directional error rate (# UMIs containing each base / # UMI families, 3 rates at each position depending on the reference base) at each position in the panel.

Blacklisting was performed using the positional UMI and R1/R2 error rates derived from the healthy samples, as they are expected to only arise from technical noise. If either of these errors was below a threshold determined from the background error rate for each type, then the location was “error blacklisted”. The thresholds used were 1% for the UMI error rate and 0.5% for the R1/R2 error rate, and an additional constraint was implemented of a maximum of a maximum of 10,000 panel positions blacklisted by error type.

Of all the variants identified by at least one variant caller, two groups were distinguished:

1. high-confidence variants, called by at least three variant callers (3+ variants), and (2) medium-confidence variants, called by at least two variant callers (2+ variants). All remaining variants were discarded.

The following filters were applied to all candidate variants, regardless of their confidence level:

1. Frequency in the panel of normal – variants identified in more than 1 normal sample (FRACTION = 0.02) were discarded.
2. On target location – only variant overlapping the capture panel coordinate were kept.
3. Overlap with blacklisted locations – regardless of the confidence level, variant in error-prone locations were not considered further.

The following additional filters were applied to the medium-confidence variants:

1. Germline support – variants were discarded if more than 1 supporting read was found in the matched germline sample.
2. High error probability – we used the binomial distribution to assess the likelihood of encountering the number of errors that could resemble the observed variant. We assumed that each sequencing event is an independent Bernoulli trial with a fixed error probability. For a given number of trials *n* (sequencing depth at the variant loci) and a per-trial error probability *p* (estimated directional error rate from the cfDNA of healthy donors given the genomic location and specific base change), the probability of observing exactly *i* errors (allele depth) is defined by:

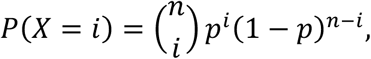

To determine the probability of having at least *k* errors, we first compute the cumulative probability of observing fewer than *k* errors:

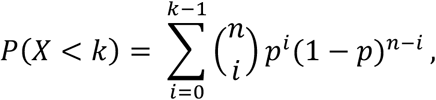

and then take the complement to quantify the probability of encountering at least *k* errors:

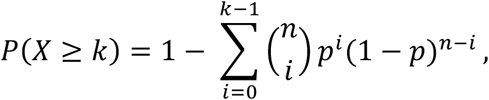

Variants were filtered if the computed probability of error was larger than 0.0001.

1. 3. UMI family support - Each variant had to be supported by at least 10 UMI families of size two or greater, to minimize the impact of PCR duplicates and sequencing errors.
2. 4. Strand bias - A Fisher exact test was performed to determine whether the strand distribution of reads supporting the variant was similar to the balance in the reference allele. Variants with significantly different strand composition (Fisher test p-value < 0.01) were removed.

Only variants passing all the filters specified were considered further.

The filtering approach was validated on the healthy samples. All potential SNVs were identified using bcftools mpileup on the aligned bam files and any variants with VAF above 5% were removed to avoid SNPs and simulate a low confidence variant calling set. The numbers of reference and variant supporting reads were then summarized by the trinucleotide base context at each panel position, using the most commonly occurring variant for positions with multiple alternative alleles. All filters applied to the 2+ plasma variants were then applied individually and in combination, resulting in a synergistic drop in error rate of ∼100-fold when all filters were applied (**Fig. S3 D, E**).

#### Variant annotation and driver mutation identification

##### LCR-BCCRC modules: vcf2maf, aule_out_tables

Retained SNVs were annotated with vcf2maf and ENSEMBL Variant Effect Predictor (version 105). ENSEMBL canonical transcripts were selected for variant consequence prediction. Mutations were evaluated and assigned a binary driver status (1 for driver; 0 for non-driver) according to the following criteria:

- Tumor Suppressor Gene (TSG) alterations: Mutations affecting genes in a curated list of tumor suppressor genes were considered driver events if the mutational consequence was among a set of defined deleterious alterations. In addition, for

mutations in TSGs that affected splice sites (classified as Splice_Site), a driver annotation was assigned.

- Whitelist variants: Independently of gene or consequence, mutations flagged in a whitelist, which contains recurrently mutated loci reported in previous publications^4,5,7^ were designated as driver events.
- Gene-specific criteria:
- `o` *NOTCH1* and *NOTCH2*: mutations that occurred in exon 34 and had consequences consistent with deleterious effects (as per TSG annotation) were considered drivers. Mutations in the 3’ untranslated region (3’UTR) of *NOTCH1* were also flagged as drivers.
- `o` *NFKBIZ*: mutations with a start position within the defined genomic range in hg38 (chromosome 3, positions 101859371 to 101859522) were annotated as drivers.
- `o` To identify pathogenic *TP53* variants we used UMD TP53 variant database (https://p53.fr/tp53-database), which includes the largest collection of curated *TP53* variants. Driver mutations were identified as follows: *TP53* mutations classified as “Pathogenic” or “Likely Pathogenic” in the UMD TP53 database, OR, all truncating mutations annotated by VEP as frameshift, splice site, in-frame deletions or insertions or events greater than 20 bases in length.

Any mutation not meeting the above conditions was classified as a non-driver event.

#### Calculation of SNV-based ctDNA burden

The SNV-based ctDNA burden for each patient was calculated by multiplying the cfDNA concentration in he/ml with the mean VAF of all mutations identified in the plasma that passed our filters.

An analysis based on Maximally Selected Rank and Statistics^24^ was carried out to determine the optimal ctDNA burden threshold to separate patients into two treatment response groups. Thresholds of 1.6 and 3.3 (log10) hGE/ml were chosen to maximize the standardized log-rank statistic using 1000-fold bootstrapping to determine the confidence intervals.

#### Molecular subtyping

##### AULE module: lymphgen_subtyping

We used a browser tool <u>LymphGen 2.0</u> to perform the molecular subtyping^6^. Input files (sample annotation file, mutation flat file and mutation gene list file) were automatically generated from a combined MAF files as specified by the developer. The chromosome location of the start of the mutation were provided in hg19 coordinates. The genomic coordinates were cross-mapped from hg38 using LiftOver function from rtracklayer and hg38ToHg19.over.chain.gz chain.

For composite cases not assigned to a single subtype, we re-evaluated subtype-specific confidence scores from the LymphGen algorithm. When a single maximum score could be identified, the sample was reclassified as “[dominant_subtype]-COMP” (e.g., “MCD- COMP” for a dominant MCD subtype), retaining its composite identity; in a simplified subtyping scheme for clinical use, the “-COMP” suffix was dropped, assigning only the dominant subtype (e.g., “MCD”).

#### Identification of inadequate plasma

Samples with fewer than 25 called SNVs were considered inadequate for accurate genotyping. We evaluated a series of pre-sequencing factors - namely, age, sex, Ann- Arbour stage, the presence of more than one extranodal site, an ECOG performance status of at least 2, elevated LDH levels, sample collection site, cfDNA concentration (measured before library preparation), and TMTV - to assess whether inadequate plasma samples can be predicted prior to sequencing completion. Cases were divided into training and test set with a split ratio of 0.75. Cases with missing TMTV values were excluded. Feature selection was performed using 2 metrics evaluating the predictive relevance of each feature for binary classification task (plasma adequate or inadequate). We used an importance-based filter leveraging a random forest learner to assess each feature’s contribution to reducing prediction error and an information gain score to quantify the amount of information each feature provides about the target variable. Top 5 features with the highest scores combined were selected for further training. Logistic regression, random forest, and naïve Bayes classifiers were trained on a designated training subset. Five-fold cross-validation was employed to assess model performance. Hyperparameter tuning was performed for the random forest classifier via grid search over a predefined parameter space - including the number of trees (ranging from 100 to 1000), the mtry ratio (from 0 to 1), and the sample fraction (from 0.01 to 1) - with classification error as the tuning criterion. The optimized random forest model was subsequently re-trained on the full training set. Partial dependence plots were generated from the random forest model to visualize the marginal effect of individual predictors on the probability of successful genotyping. For the naïve Bayes classifier, continuous predictors were dichotomized based on empirically determined thresholds, and pairwise chi-square tests were conducted to confirm the independence of the binary variables. The final models were then evaluated on a held-out test set (**Fig. S9E-H**).

The final naïve Bayes model has been implemented in an Excel-based tool designed for clinical decision-makers (**Extended Data Table 6).** This tool enables the prediction of whether a patient is likely to benefit from cfDNA-based genotyping and provides guidance on whether adjustments to sequencing depth or input DNA quantities may be necessary.

#### Structural variant calling

##### AULE module: sv_merge

SVs were identified using two complementary callers: GRIDSS and Manta with default settings. A custom R script was employed to merge the outputs from GRIDSS and Manta. For downstream analysis, only variants with a minimum of five reads supporting the break point, including at least 1 split read (SR) and at least 1 discordant read pair (RP) were retained: SR + RP >= 5 & (SR >=1 RP >=4 | SR >= 4 RP >=1). The output from the two callers was then intersected using the StructuralVariantAnnotation package. SVs partners were annotated with co-ordinates defining regions of interest (*BCL2*, *BCL6* and *MYC*). Regions of interest (*BCL2*, *BCL6*, *MYC*, IgH, IgK, IgL) were annotated using the hg38 converted co-ordinates from Chong et al (Blood Advances 2018), expanded to include the DIRECT panel. Rearrangement partners falling outside these regions of interest were labelled “Partner unknown”.

#### Copy number aberration calling

##### AULE module: ichorcna

Tumor fractions were estimated using ichorCNA pipeline, a tool designed specifically for shallow Whole Genome Sequencing (sWGS) data derived from cfDNA. Briefly, genome was partitioned into fixed-size bins (1) and read counts per bin were normalized to correct for GC-content and mappability biases. The normalized bin counts served as input for ichorCNA, which employs a hidden Markov model to segment the genome into regions of copy number alteration. A customized panel of normals (PON) was generated from 51 healthy donor samples sequenced using the same capture panel. ichorCNA estimated tumor fraction, defined as the proportion of circulating tumor DNA (ctDNA) within the cfDNA sample, were used as ctDNA tumor burden proxy. Capture-panel derived fractions were compared against sWGS samples prepared form the same samples achieving high correlation (r = 0.93, **Fig. S7B**).

#### Phased variants analysis

##### AULE modules: aule_pv, aule_pv_pon

Following the approach outlined by Kurtz et al^14^, we developed an automated phased variant detection pipeline implemented in Snakemake as Python and R scripts. The workflow takes UMI-collapsed query name-sorted BAM files as input.

##### Identification of candidate phased variants

Briefly, reads were processed as pairs, and only those meeting a user-specified minimum mapping quality threshold (MAPQ > 20 in this study) were retained for analysis. For each proper read pair, single nucleotide substitutions with a minimum base quality of 20 were identified. Discordant bases between read 1 (R1) and read 2 (R2) were discarded. Read-level metrics, such as insert size, mapping quality and UMI-family size were retained. Candidate phased variants were identified by generating all possible doublet (pair) and triplet (trios) combinations from high-quality, non-reference position within each read pair. For instance, if a read pair had 4 non-reference loci, 6 doublets and 4 triplets were considered independently. The Python package *pysam* was used to process sequencing reads and extract the necessary read or base-level information.

Candidate phased variants were collapsed by genomic regions. Allele depth was calculated as the total number of reads supporting all individual components of a phased variant. Total depth was computed as the total number of reads covering a phased variant locus. Phased variant allele frequencies (PVAFs) were calculated by dividing the number of unique deduplicated read pairs supporting a variant by the total number of unique deduplicated read pairs covering the phased variant genomic region. Individual components of identified phased variants were annotated with the gnomAD database frequencies to account for common population SNPs, and with the read support in matched germline sample.

This approach generated a list of candidate PVs, each supported by a minimum read depth of 1 in every tumor sample.

##### Genotyping of tumor-specific phased variants

We considered all available pre-treatment samples (plasma and biopsy, if available) for tumor specific genotyping. Genotyping for plasma and biopsy samples was performed independently.

We generated 2 panel of normals (PONs) from 40 healthy cfDNA donors to increase tumor specificity of identified phased variants.

1. SNV PON – generated with CreateSomaticPanelOfNormals identifying individual loci recurrently altered in healthy cohort.
2. Phased variant PON – containing all identified phased variants called as described above, except matched germline read support.

The candidate phased variants were filtered based on the following criteria:

1. Zero read support in the matched germline – all phased variant loci were required to have 0 read support in the matched germline.
2. GnomAD population frequency of individual loci less than 0.002.
3. Individual loci not listed in the SNV PON
4. Individual loci not listed in phased variant PON in more than 1 patient.
5. PVAF >= 0.002
6. At least one locus on target (within the capture panel genomic coordinates)
7. Minimum depth of the phased variant locus >= 100 read-pairs

This filtering process produced PV lists that were subsequently used for disease monitoring and MRD detection.

##### MRD test

For MRD detection, leveraging prior knowledge of the pre-treatment tumor genotype, we evaluated MRD assessment sample for the presence of any PVs identified in the pre-treatment genotyping sample. When both biopsy and plasma genotyping was available, independent MRD evaluation was performed. The MRD evaluation was implemented as in Kurtz et al. 2021^14^ .

1. Identification of genotyped PVs in the assessment sample

Briefly, starting with a list of *k* tumor-derived PVs from the pre-treatment genotyping sample, we identified all read pairs covering at least one of these PVs. We denote the total number of such read pairs (informative reads) as *d*, which represents the cumulative informative depth. We then determined how many of these d read pairs contained >= 1 of the *k* tumor-derived PVs; this number, *x*, represents the tumor-derived molecules harboring somatic PVs in the sample. The combined PVAF is calculated as the ratio x/d. PVAF was computed independently for doublet and triplet PVs in each sample.

1. 2. Determining the significance of the tumor-specific signal with Monte Carlo simulation To determine the statistical significance of detecting tumor-derived PVs in each sample, we implemented an empirical significance testing framework utilizing a Monte Carlo simulation as detailed in Kurtz et al^14^. We defined a test statistic

representing the arithmetic mean allele fraction (*PVAF_mean*) of *k* tumor-derived PVs identified in the genotyping sample. To assess whether the test statistics significantly exceeds the background error rate of comparable PVs within the same sample, we employed the following Monte Carlo approach:

1. For each of the *k* PVs, we identified 50 alternate PVs with identical base change types (e.g., A>T and C>A) and inter-variant distances, located on the same

chromosome; alternate positions with allele fractions >5% (indicative of germline SNPs) were excluded.

1. 2. From these alternates, we generated 10,000 random permutations, each comprising one alternate PV per original PV, and computed the corresponding alternate phased-variant fraction.
2. An empirical P-value was then calculated as the fraction of permutations where the observed *PVAF_mean* was less than or equal to the mean fraction from the 10,000 alternative (not tumor-specific) runs.
3. Because this process was conducted separately for doublets and triplets, the resulting P-values were combined using Fisher’s method to produce a composite significance of tumor content.

##### Selection of best genotyping specimen

Across available genotyping samples (pre-treatment plasma and/or biopsy), the best genotyping set was selected based on the highest total number of informative reads in a pre-treatment sample. Doublets and triplets were considered together. The MRD assessment sample, genotyped using this optimal genotyping set, was designated for the final MRD assessment.

##### Computing theoretical limit of detection

The limit of detection (LoD) represents the minimum tumor fraction in a sample that can be theoretically distinguished from background noise with a specified sensitivity and false-positive rate. We utilized a binomial detection model, where this minimal tumor fraction (LoD) is inferred from the probability of detecting variants given the number of informative reads (IR), expected detection probability and false positive rate. We defined the desired detection probability (*P_detection*), typically 0.95 for 95% sensitivity, and a false-positive rate (α = 0.05), then adjusted the non-detection probability as:

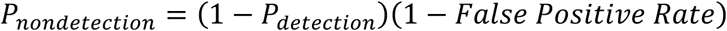

The LoD was then calculated:

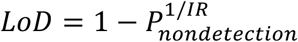

This equation modelled the exponential decrease in non-detection probability with increasing IR, yielding the minimum theoretical tumor fraction detectable under the given conditions.

#### Library saturation analysis

To quantitatively assess improvements in cohort-wide sensitivity from increasing either DNA input or sequencing depth, we performed a saturation analysis. We assumed that the discovery of new DNA molecules follows a smooth saturating process (all molecules can be sequenced by the same diminishing-return dynamics), that there is a fixed total number of unique molecules in a sample, and that new molecules are sampled with uniform efficiency. The theoretical maximum number of unique molecules recoverable, Umax, was represented as:

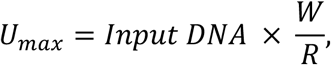

Where *W* denotes the total cumulative width of the regions with tracked variants, and *R* denotes the average read length. Input DNA was represented in haploid genome equivalents, hGE (input mass in ng x 1000/3.3). Comparable estimates of the total number of unique reads in a sample were obtained using empirical Chao1 or Good-Turing estimators.

The relationship between the number of unique molecules and the number of sequenced reads, *U(N),* was modelled with Michaelis-Menten equation, where:

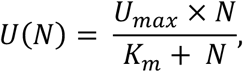

where *N* is the number of molecules sequenced and *Km* represents the number of reads needed to reach half of *Umax*. *Km* was derived empirically from the data. For a cohort-wide analysis **(Fig 5I)** we used the average number of unique molecules corresponding to average number of sequenced reads. Three input DNA scenario (25ng, 50ng and 75ng) were considered. This analysis was performed under the naïve assumption that the ratio of unique to sequenced molecules remains constant across different input amounts. The estimated number of unique molecules (informative reads) was compared to the theoretical limit of detection computed as described above.

### Statistical analysis

In this translational, feasibility study, there was no research treatment and patients received standard of care immunochemotherapy. The primary objective was to establish a robust molecular pipeline to identify trackable mutations in ctDNA from patients undergoing treatment for high-grade B cell lymphoma. The clinical secondary outcomes included: (1) Assess the utility of serial ctDNA assessment as a predicator of clinical outcome in high-grade B cell lymphoma. (2) Assess the utility of integrated data from clinical risk factors (IPI), up-front genotype, serial ctDNA response and radiological assessment (CT or PET-CT). The clinical outcomes include end of treatment response, TTP, PFS and OS as described in the previous section.

Due to the exploratory nature of this study, there was no formal sample size calculation associated with the number of participants. However, the main aim of the study was to establish a molecular pipeline, and to find trackable mutations in 75%. The proposed sample size of 150 evaluable patients was more than adequate to account for a significance level of 0.05 with 90% power.

At the end of stage 1 the first 50 patients enrolled into the study were assessed for trackable mutations with the following plan. If at least 30 participants yielded successful identification of trackable mutations, then the study would proceed to stage 2 as is. If not, the method for identifying trackable mutations would be amended to satisfy the feasibility criterion. The number 30 stems from the lower bound of a Clopper-Pearson exact confidence interval based on 50 patients and 75% probability. In the interim analysis (stage I), after 50 evaluable participants were included in the study, trackable mutations were identified in 30 (90.91%, 95% CI 75.67% to 98.08%) out of 33 participants with available samples. The study continued to the second stage without any adjustment.

The full eligible population is referred to as Cohort 1 and used as primary population. A more homogeneous subgroup participants excluding participants with Burkitt lymphoma or those who received non-anthracycline-based chemotherapy is referred to as Cohort. 2. Kaplan–Meier plots were generated for time-to-event outcomes and groups were compared using the log-rank test. No multiplicity adjustments were performed. The ichorCNA fraction and SNV-derived ctDNA concentrations were correlated with time-to-event outcomes via a Cox proportional hazard regression model adjusting for IPI score as continuous and log transformed TMTV score. To explore the optimal cutoff point of the ctDNA concentration and ichorCNA fraction on TTP, we used maximally selected rank statistics method with a cut-oJ point that maximized the absolute standardized log-rank statistics^24^. All statistical analyses were carried out in R (v4.3.1) and maxstat (0.7-25), all P values are based on two-tailed tests.

## Conflicts of Interest

DJH: reports research funding from Astra Zeneca and GSK.

SB: research funding from Beigene, F Hoffman la Roche and Takeda

SKM: Speaker fees and conference travel: SOBI, Alexion-AZ

DH and VM – employment and stock ownership – AstraZeneca

## Data Availability

All sequencing files have been deposited at the European Genome Phenome Archive (EGA) with accession number: EGAS50000000968.

## Acknowledgements

We thank the patients, and the families and friends who supported them, for participating in this study. This study was sponsored by Cambridge University Hospitals NHS Foundation Trust and the University of Cambridge and financed by a grant from the Mark Foundation for Integrated Cancer Medicine to the CRUK Cancer Centre and by AstraZeneca. We thank Cambridge Clinical Trials Unit – Cancer Theme for their core staff support and the clinical trials support staff at all participating sites. This work was supported by the Cancer Molecular Diagnostics Lab at the Cancer Research UK Cambridge Centre [CTRQQR-2021\100012] and by Tissue Bank. We thank CMDL technical staff for blood processing, extractions and library prep and Katie Honan (CMDL) for sequencing data transfer. This research was supported by the NIHR Cambridge Biomedical Research Centre (NIHR203312) and NIHR Leicester Biomedical Research Centre (NIHR203327). The views expressed are those of the authors and not necessarily those of the NIHR or the Department of Health and Social Care. NHC was supported by a clinical research training fellowship from CRUK Cambridge Centre. DJH was supported by a fellowship from Cancer Research UK (CRUK; RCCFEL\100072) and received core funding from Wellcome (203151/Z/16/Z) to the Wellcome-MRC Cambridge Stem Cell Institute and from the CRUK Cambridge Centre (A25117). For the purpose of Open Access, the authors have applied a CC BY public copyright license to any Author Accepted Manuscript version arising from this submission.

## Data and code availability

All code including the end-to-end pipeline used for plasma genotyping, ctDNA burden quantification and phased variant supported MRD is made freely available under an open-source license at (https://github.com/Hodson-Bioinformatics/Aule/). All sequencing files have been deposited at the European Genome Phenome Archive (EGA) with accession number: EGAS50000000968.

## Supplementary Figure Legends

**Supplementary Figure 1.**
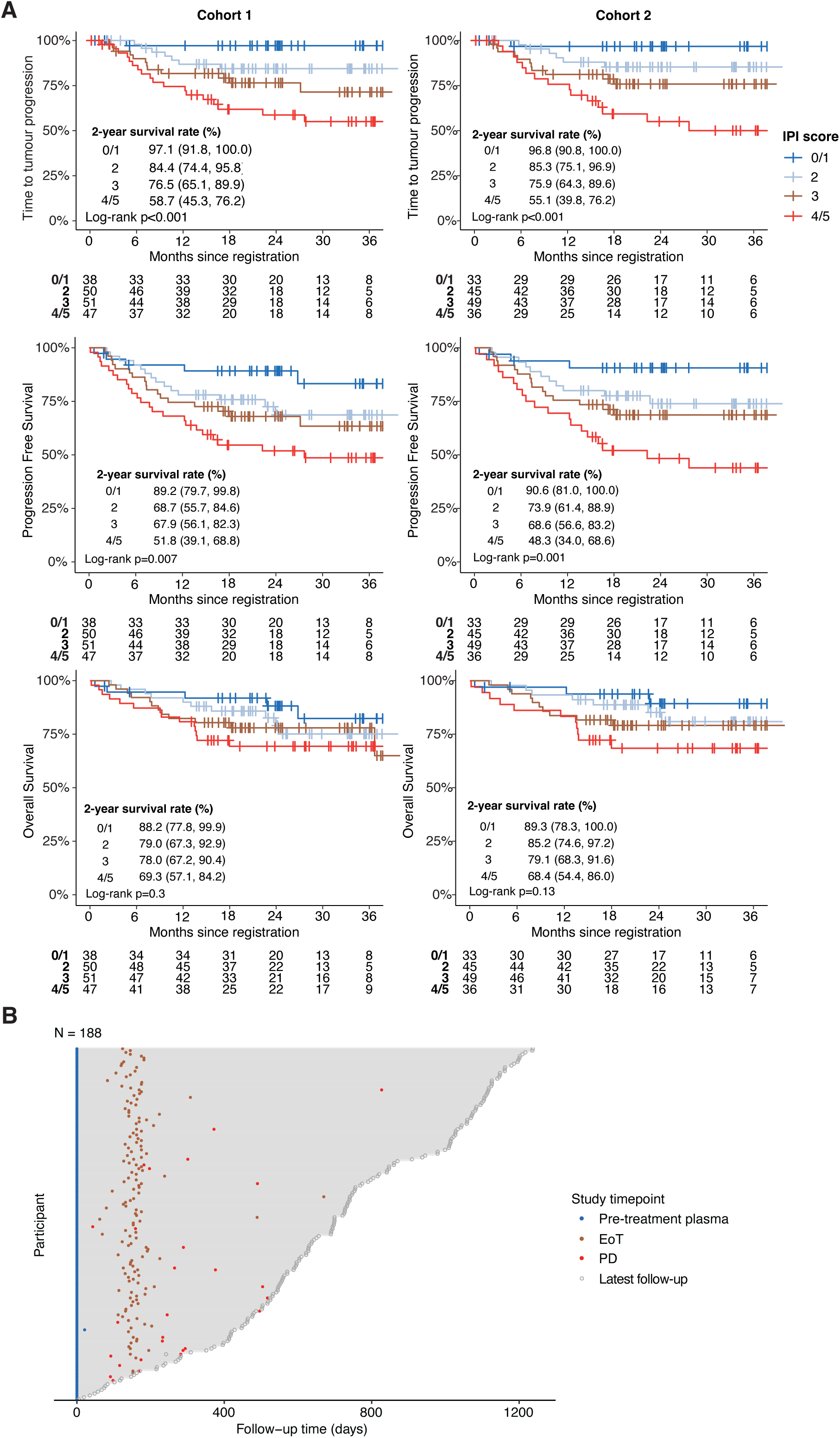
Baseline characteristics and summary of samples collected. **A**. Full clinical outcome data (TTP, PFS and OS) for all patients in Cohort 1 (lè) and Cohort 2 (right) stratified by IPI. **B.** Timings of sample collections across all patients enrolled in the DIRECT study. The survival curves are compared with a two-sided log-rank test and no multiple comparison performed.

**Supplementary Figure 2.**
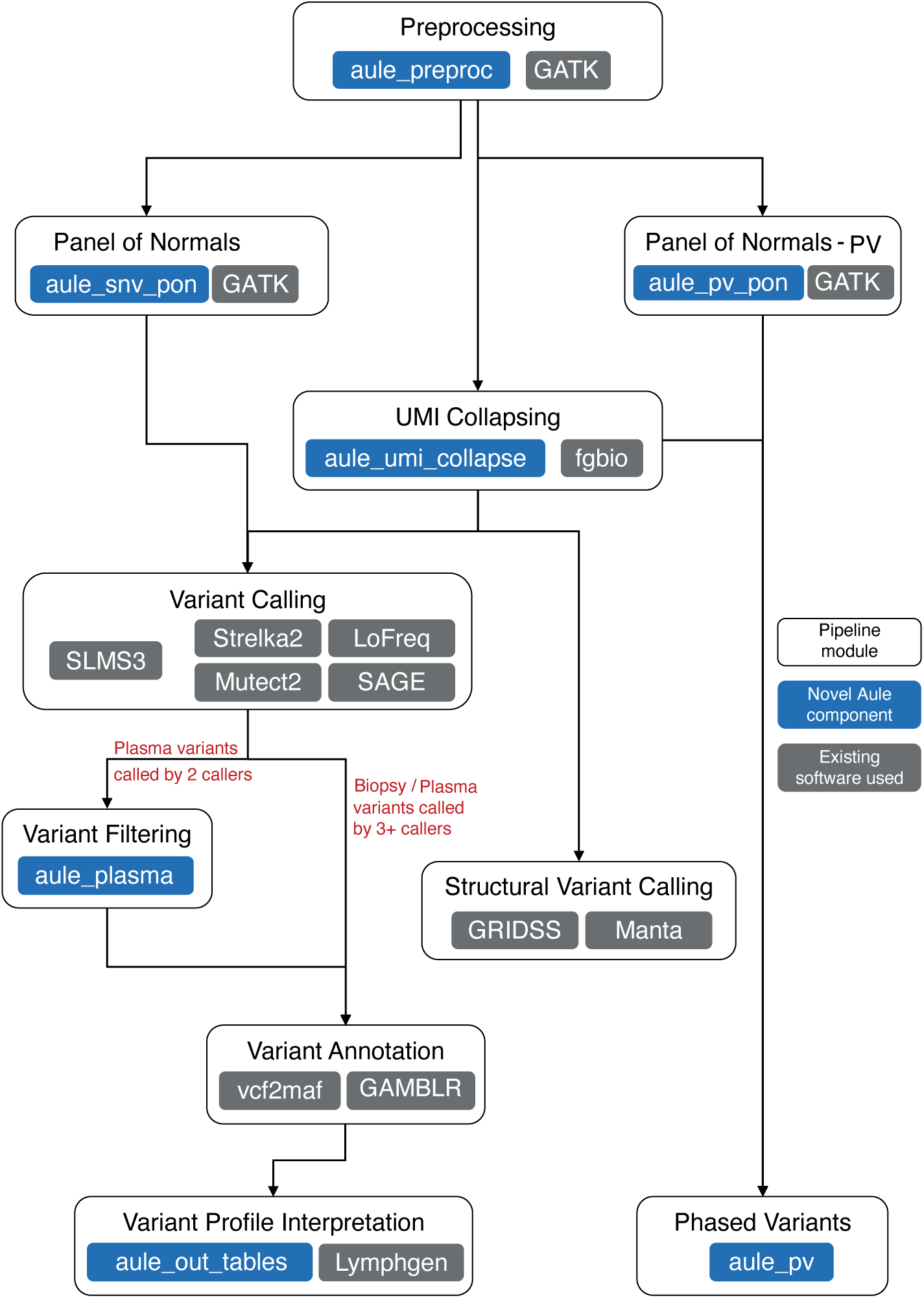
Overview of Automated Ultrasensitive Lymphoma Evaluation (AULE) bioinformatic pipeline used for processing targeted sequencing data.

**Supplementary Figure 3.**
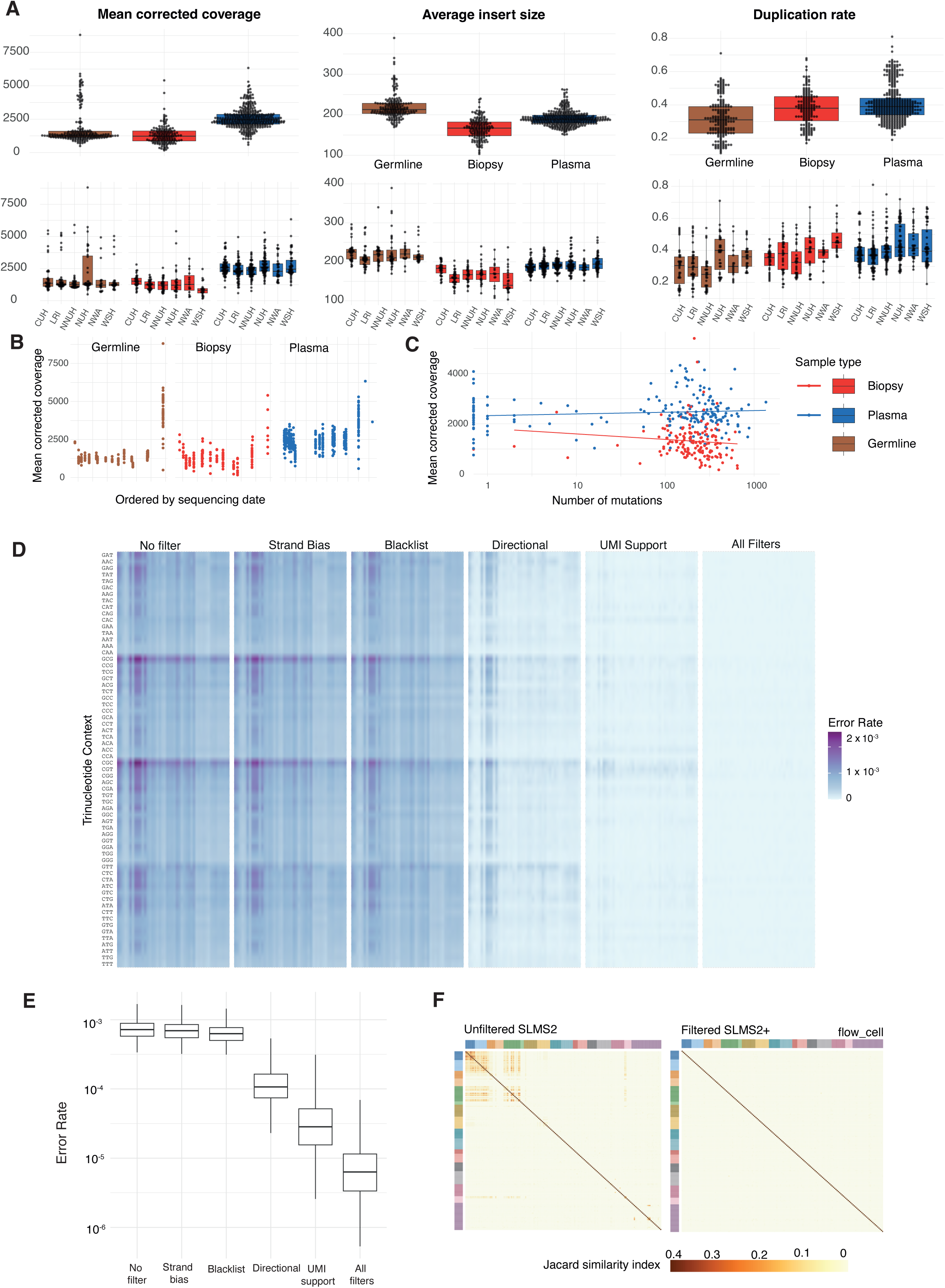
Quality control metrics and impact of error suppression strategies. **A.** QC metrics, showing Mean corrected coverage (lè), average insert size (central) and duplication rate (right) for all samples sequenced in DIRECT study. Data are split by enrolment site in lower panels. **B.** Mean corrected coverage by 6me, with cases ordered by sequencing date. **C.** Total number of variants identified is ploged against mean corrected coverage for each sample showing absence of any linear relationship between corrected coverage and variant number across the range of coverage achieved in DIRECT. **D.** Effect of the indicated customized error suppression strategies upon background noise presented in trinucleotide context, assessed across a panel of 46 normal plasma samples. **E.** Effect of indicated customized error suppression strategies upon background noise assessed, either individually or in combination, across a panel of 46 normal plasma samples. **F.** The Jaccard similarity index between patients before (lè) and after (right) customized error suppression.

**Supplementary Figure 4.**
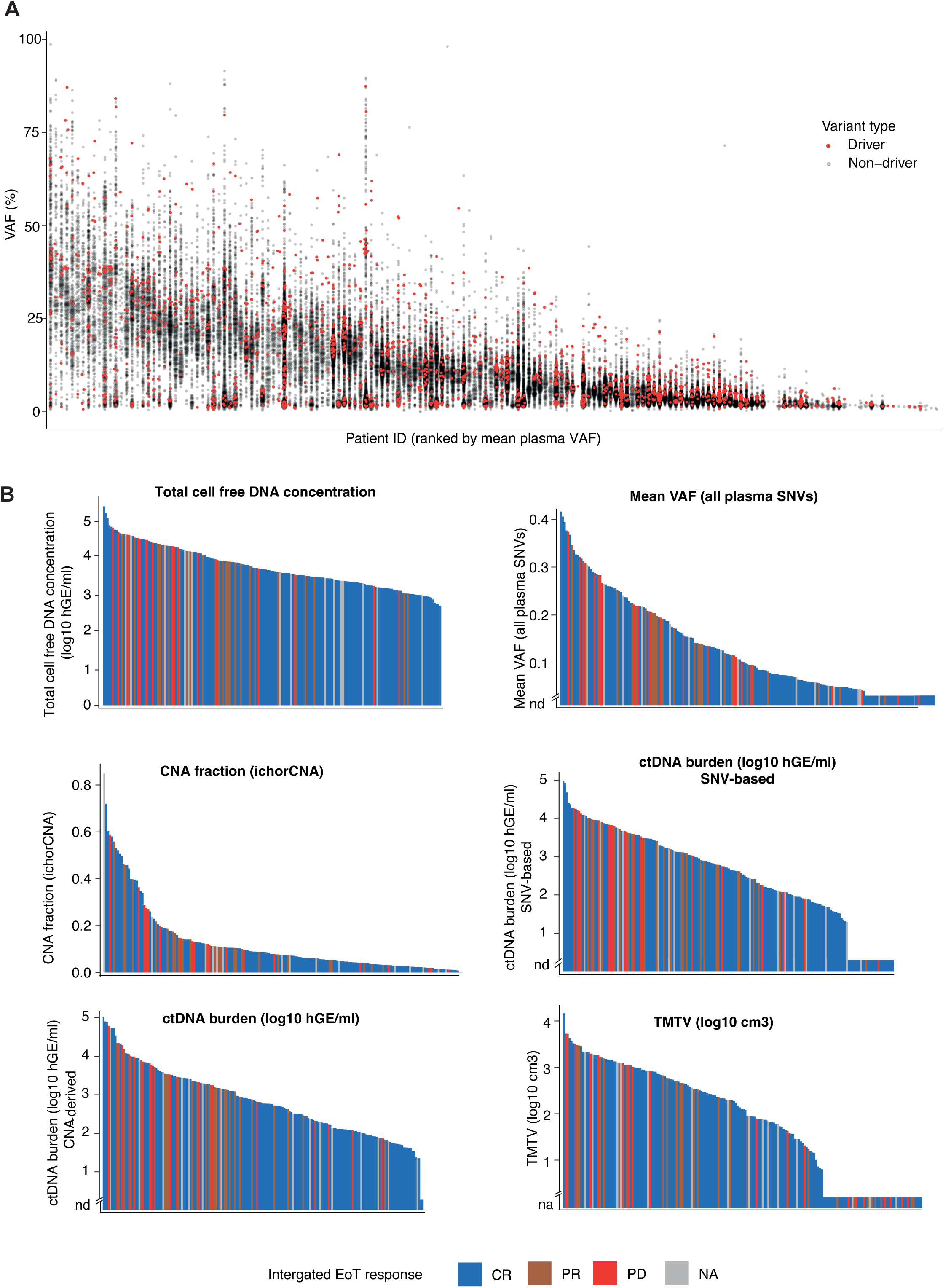
Use of pretreatment ctDNA metrics to predict clinical outcome. **A.** All 164 patients with variants detected from plasma sequencing are ranked from high to low tumor burden. The variant allele fraction for each variant is shown as a data point. Driver variants are colored in red. **B**. Bar charts showing association of pretreatment ctDNA metrics, and TMTV, with EoT radiological response assessment for all patients in Cohort 1.

**Supplementary Figure 5.**
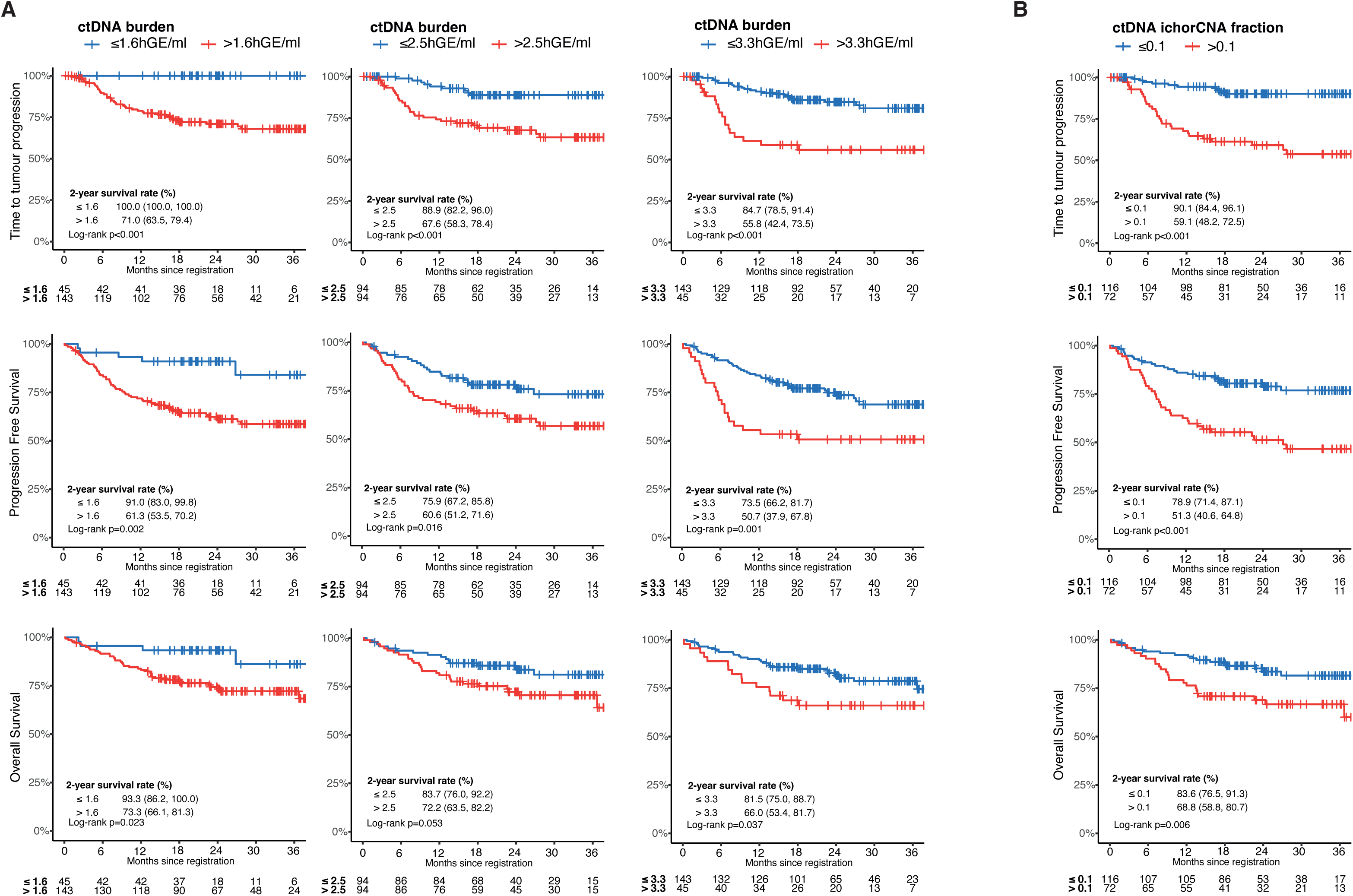
Survival outcomes of Cohort 1 by pretreatment ctDNA metrics. A. TTP, PFS and OS are shown for patients in Cohort 1 stratified by pretreatment, SNV-derived ctDNA burden using thresholds of 1.6 log10 hGE/ml (lè), 2.5 log10 hGE/ml (center) and 3.3 log10 hGE/ml (right). **B.** TTP, PFS and OS are shown for patients in Cohort 1 stratified by ichorCNA-derived tumor fraction using a threshold of 0.1. The survival curves are compared with a two-sided log-rank test.

**Supplementary Figure 6.**
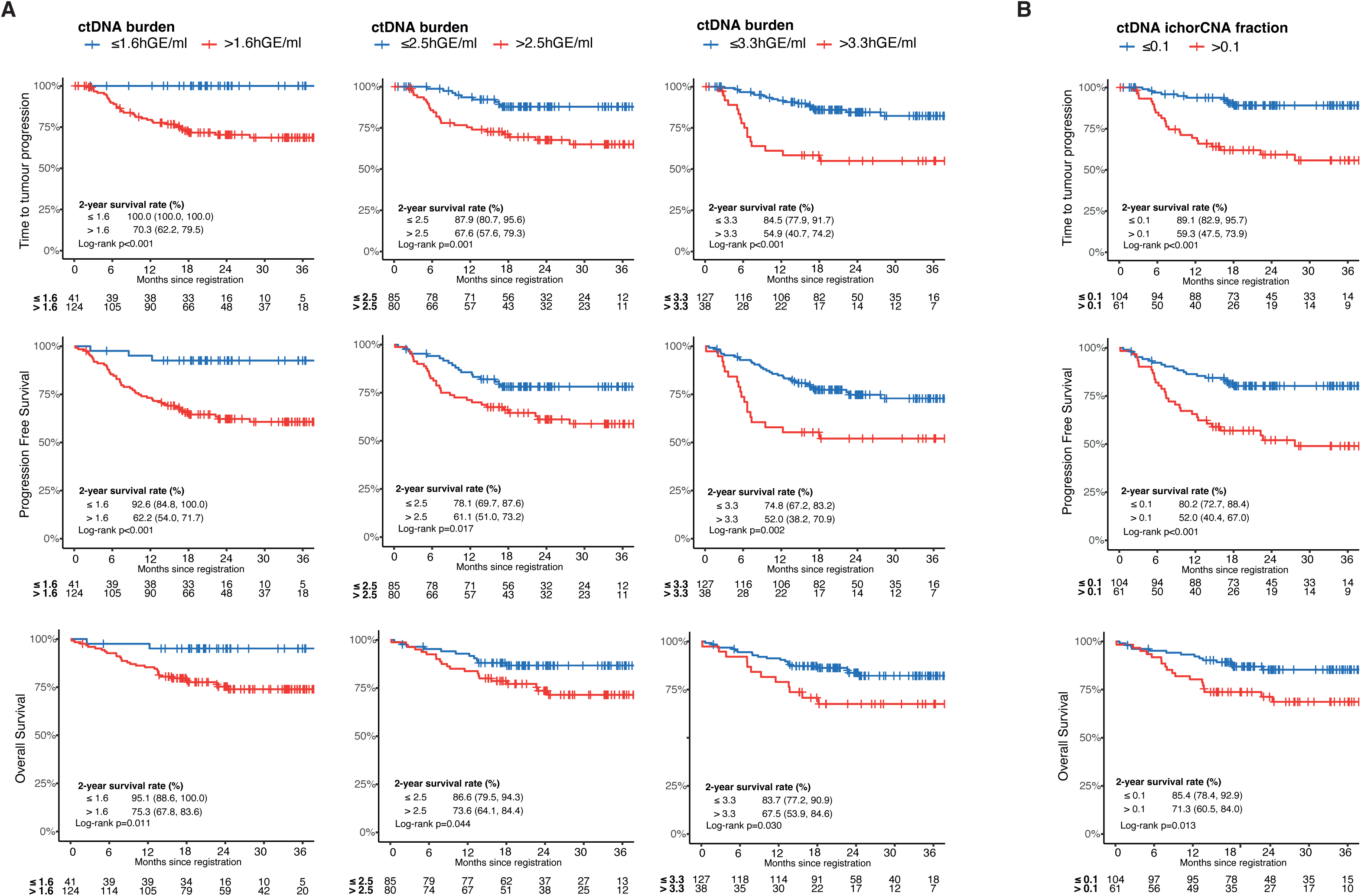
Survival outcomes of Cohort 2 by pretreatment ctDNA metrics. **A.** TTP, PFS and OS are shown for patients in Cohort 2 stratified by pretreatment, SNV-derived ctDNA burden using thresholds of 1.6 log10 hGE/ml (lè), 2.5 log10 hGE/ml (center) and 3.3 log10 hGE/ml (right). **B.** TTP, PFS and OS are shown for patients in Cohort 2 stra6fied by ichorCNA-derived tumor fraction using a threshold of 0.1. The survival curves are compared with a two-sided log-rank test.

**Supplementary Figure 7.**
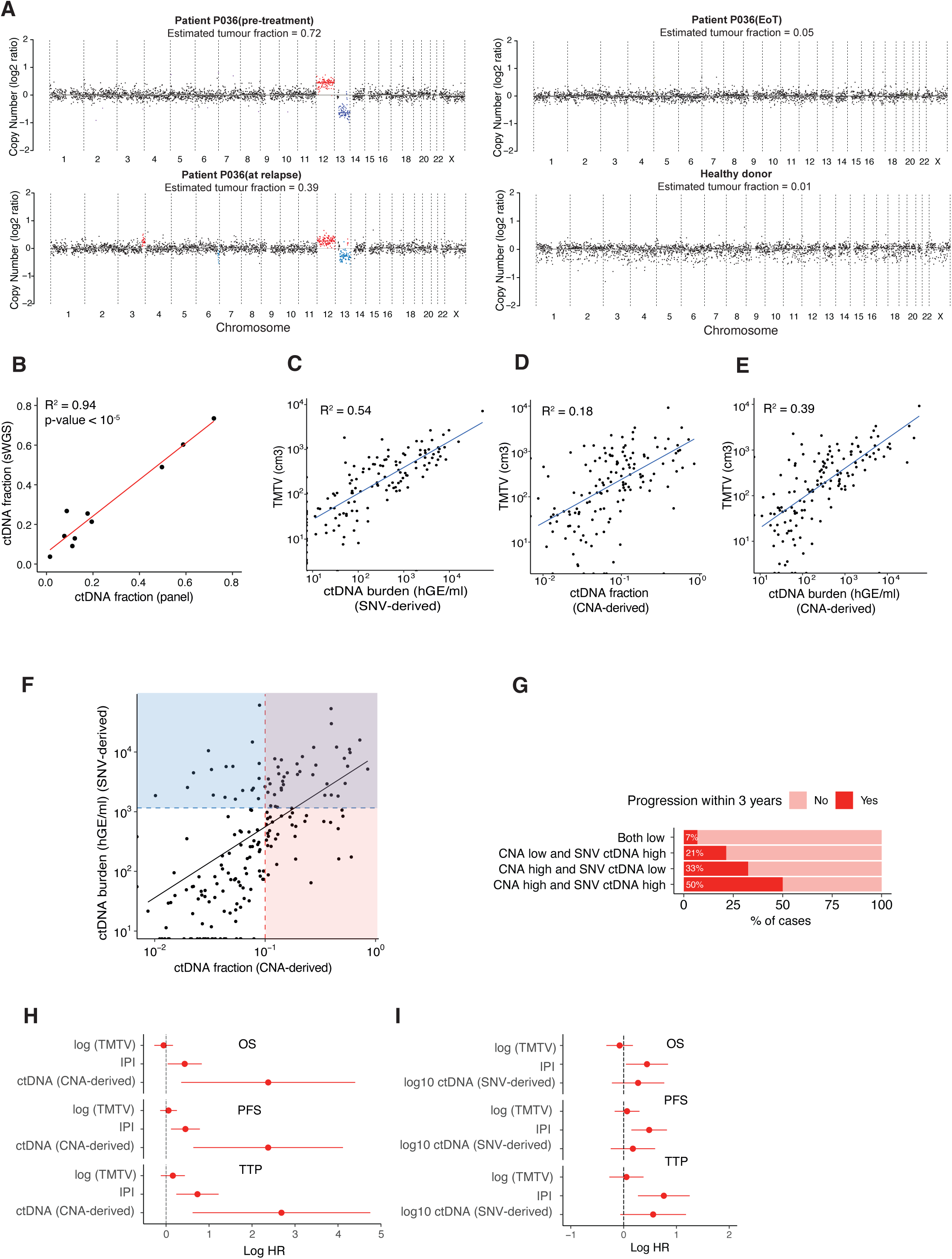
Quantification of tumor fraction from targeted panel data using ichorCNA. **A.** Example data from ichorCNA from a typical DIRECT patient at presentation, remission and relapse, and from a healthy normal donor. **B.** Comparison of ctDNA tumor fraction from matched samples when ichorCNA is applied to shallow whole genome sequencing (sWGS) or panel-based sequencing. **C-E.** Comparison PET-derived pretreatment TMTV with SNV-derived ctDNA burden (**C**), ichorCNA-derived tumor fraction (**D**), and ichorCNA-derived ctDNA burden (tumor fraction x cfDNA concentration) (**E**). **F.** Comparison of SNV-derived ctDNA burden with ichorCNA tumor fraction. R² and p-value, derived from a linear model, are shown. **G.** Outcome (progression) at 12 months for the 4 quadrants depicted in **F**. **H.** Forrest plots depicting hazard ratios from multivariable analysis of TMTV, IPI and ctDNA fraction (ichorCNA-derived) using TTP, PFS and OS as indicated. **I.** Forrest plots depicting hazard ratios from multivariable analysis of TMTV, IPI and ctDNA burden (SNV-derived) using TTP, PFS and OS as indicated. Risk factors were considered as continuous variables.

**Supplementary Figure 8.**
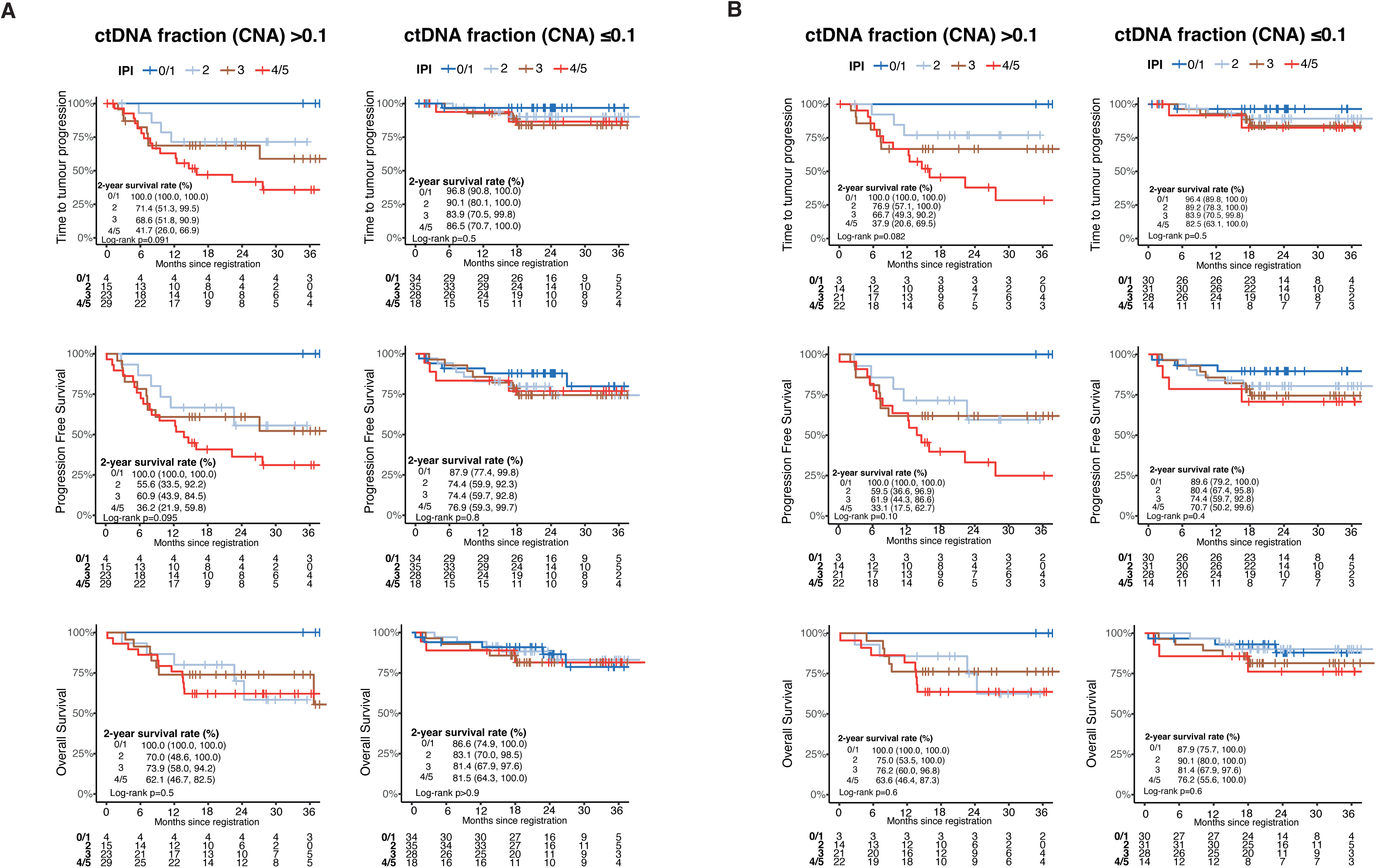
Survival by ctDNA fraction and IPI. TTP, PFS and OS are shown for cases with high (>0.1) (lè) and low (≤0.1) (right) pretreatment ctDNA fraction derived from ichorCNA, stratified by IPI score. **A.** Shows outcomes for Cohort 1. **B.** Shows outcomes for Cohort 2. The survival curves are compared with a two-sided log-rank test and no multiple comparison performed.

**Supplementary Figure 9.**
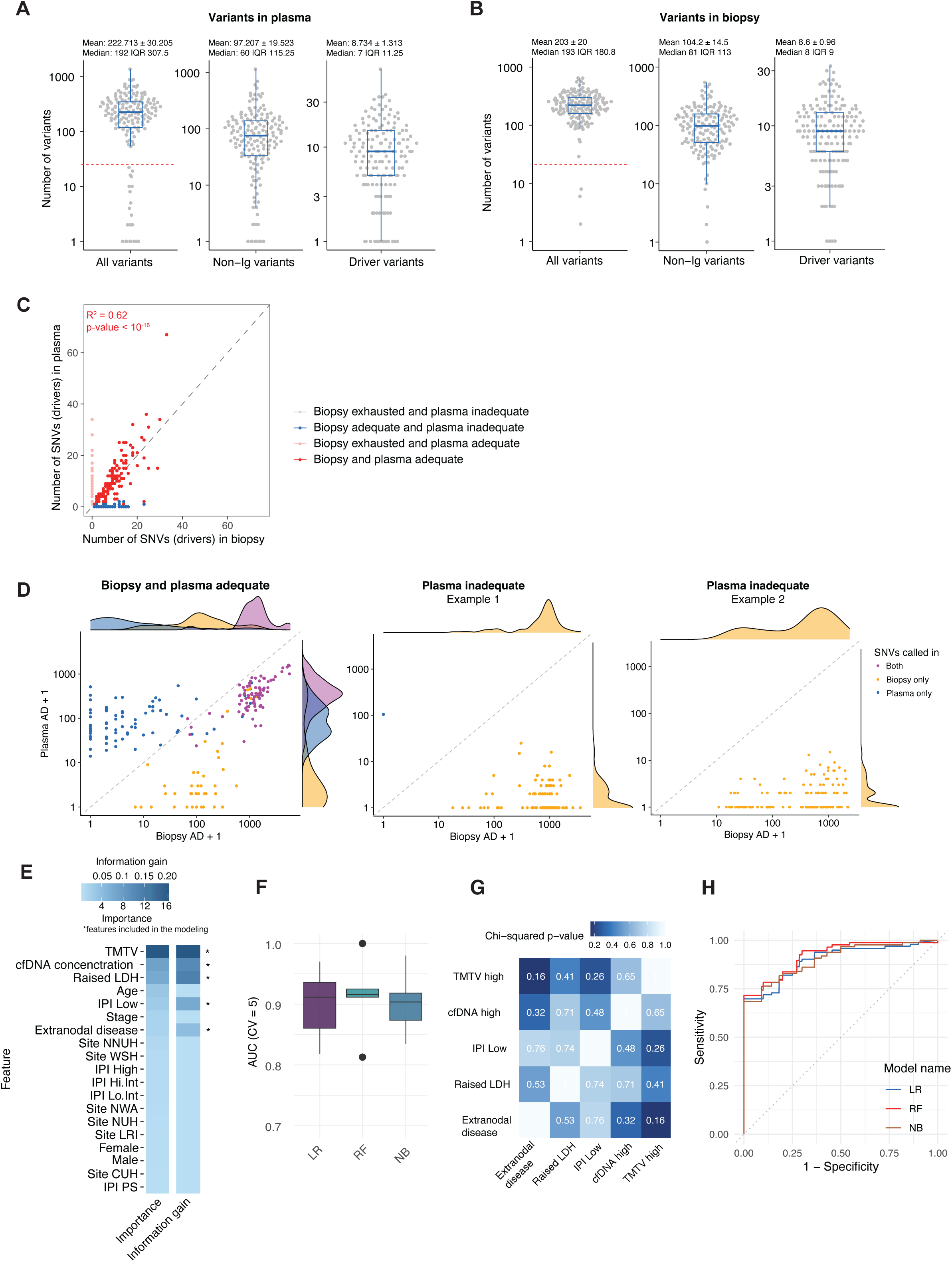
Predictors of success and failure for plasma genotyping. **A.** Number of variants (all variants, non-Ig variants and driver variants) detected in plasma sequencing showing the QC threshold of 25 total variants used to define failed samples (dashed red line). **B.** Total number of variants identified in biopsy samples. **C**. Total number of driver mutations comparing matched plasma and biopsy samples. Samples are colored by whether they passed QC thresholds for plasma and biopsy genotyping. **D.** Forced calling of all SNVs called in each case was performed on plasma and biopsy samples to identify cause for missing variants. Lè panel shows a case adequately genotyped by both plasma and biopsy. Right panels show two representative cases where plasma genotyping was inadequate, showing absence or very low levels of the mutant reads within the plasma libraries. **E.** Heatmap showing feature importance and information gain for pre-sequencing factors predicting success of failure of plasma-based genotyping. **F.** Overview of the five-fold cross- validation training set results for three models—logistic regression (LR), random forest (RF), and naïve Bayes (NB)—evaluated for predicting plasma-based genotyping success, with performance measured by the area under the curve (AUC). **G.** Heatmap illustrating the p- values from a chi-square test of independence among the selected predictors. **H.** Test set performance for three models evaluated for predicting plasma-based genotyping success.

**Supplementary Figure 10.**
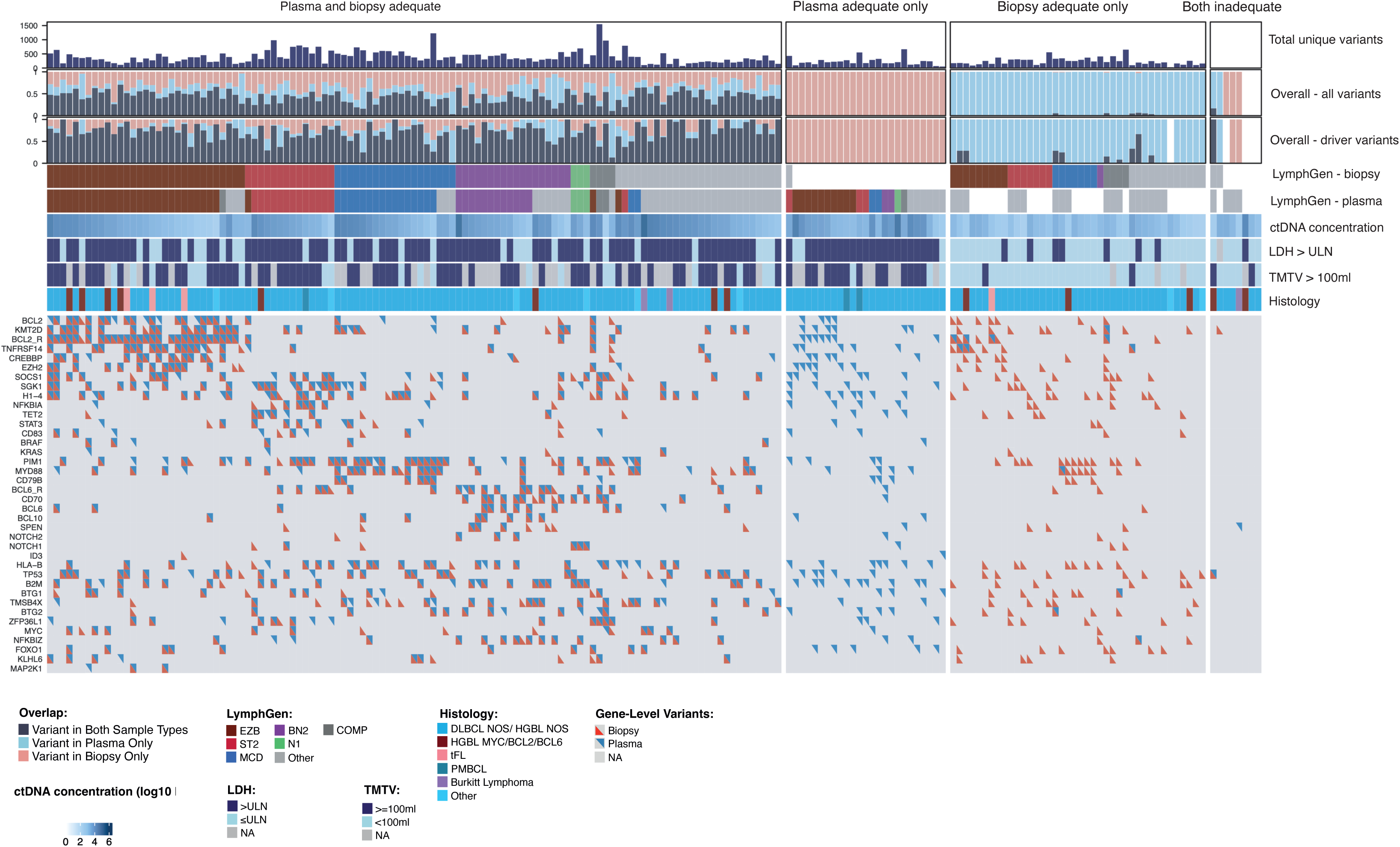
Summary of sequencing data for all DIRECT patients. Oncoplot comparing outcome of plasma and biopsy genotyping for the 30 most commonly mutated genes across all samples.

**Supplementary Figure 11.**
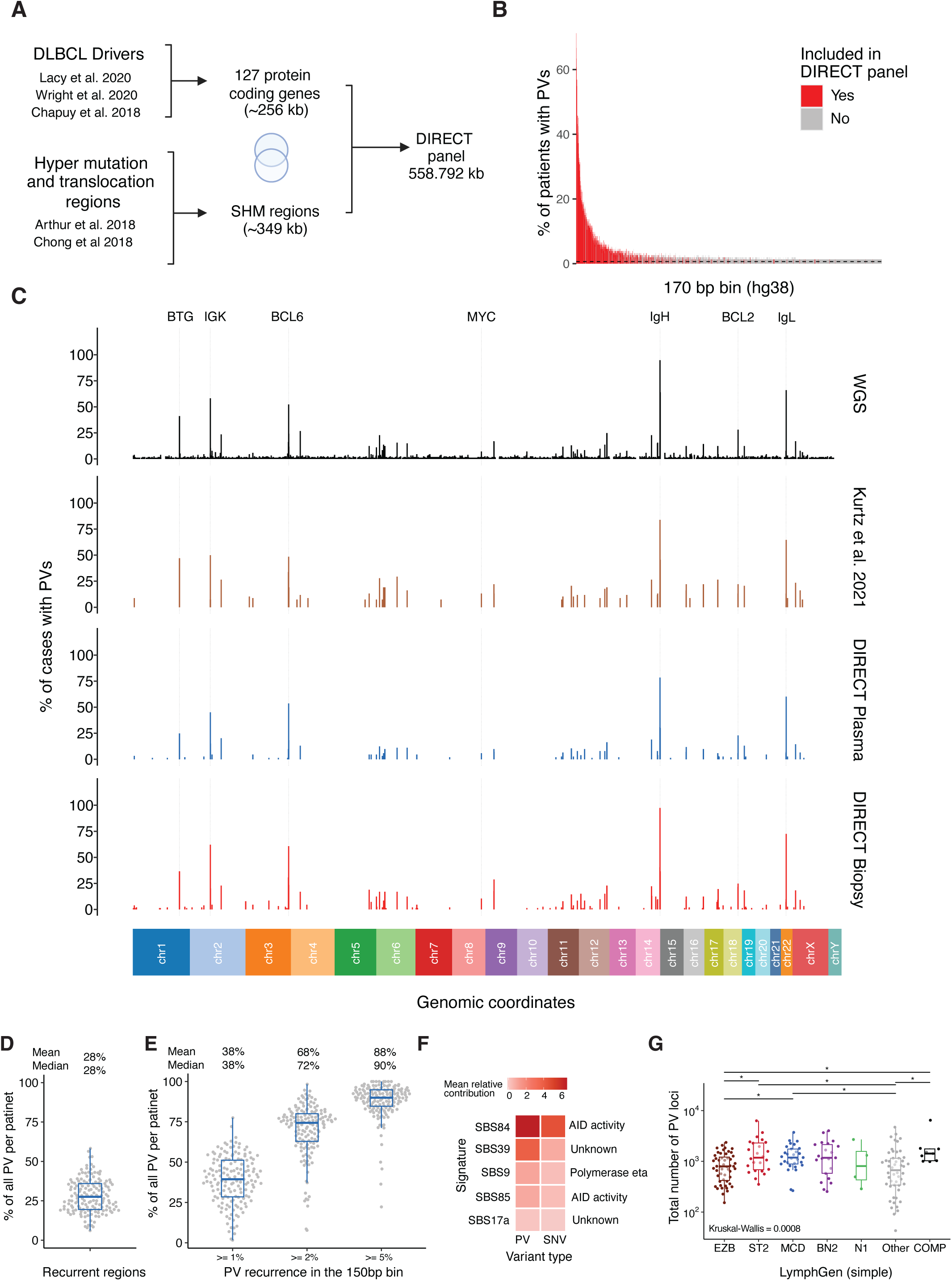
Design of a targeted sequencing panel for phased variant supported MRD assessment. **A.** Flowchart showing target region selection in the DIRECT sequencing panel including recurrent DLBCL drivers with somatic hypermutations. **B.** Bar plot illustrating the recurrence of candidate phased variants (PVs) across genomic regions in whole-genome sequencing (WGS) data^23^, aggregated into 170 bp bins, with regions in the DIRECT panel highlighted in red. **C.** Bar plots showing the distribution of phased variants (PVs) across 1,000-bp genomic bins in published studies versus targeted sequencing of plasma and biopsy in DIRECT cases. **D.** Proportion of all PVs from WGS (Arthur et al 2018) falling within 150bp bins that recurrently contain PVs in at least 1% of patients. **E.** Proportion of all PVs from WGS (Arthur et al 2018) falling within regions included in the design of our panel. **F.** Heatmap showing the top 5 mutational signatures enriched in identified phased variants (PVs) compared to SNVs. **F.** Boxplot showing the total number of phased variants (PVs) loci by genetic subtype. * p < 0.05, pairwise Wilcoxon test.

**Supplementary Figure 12.**
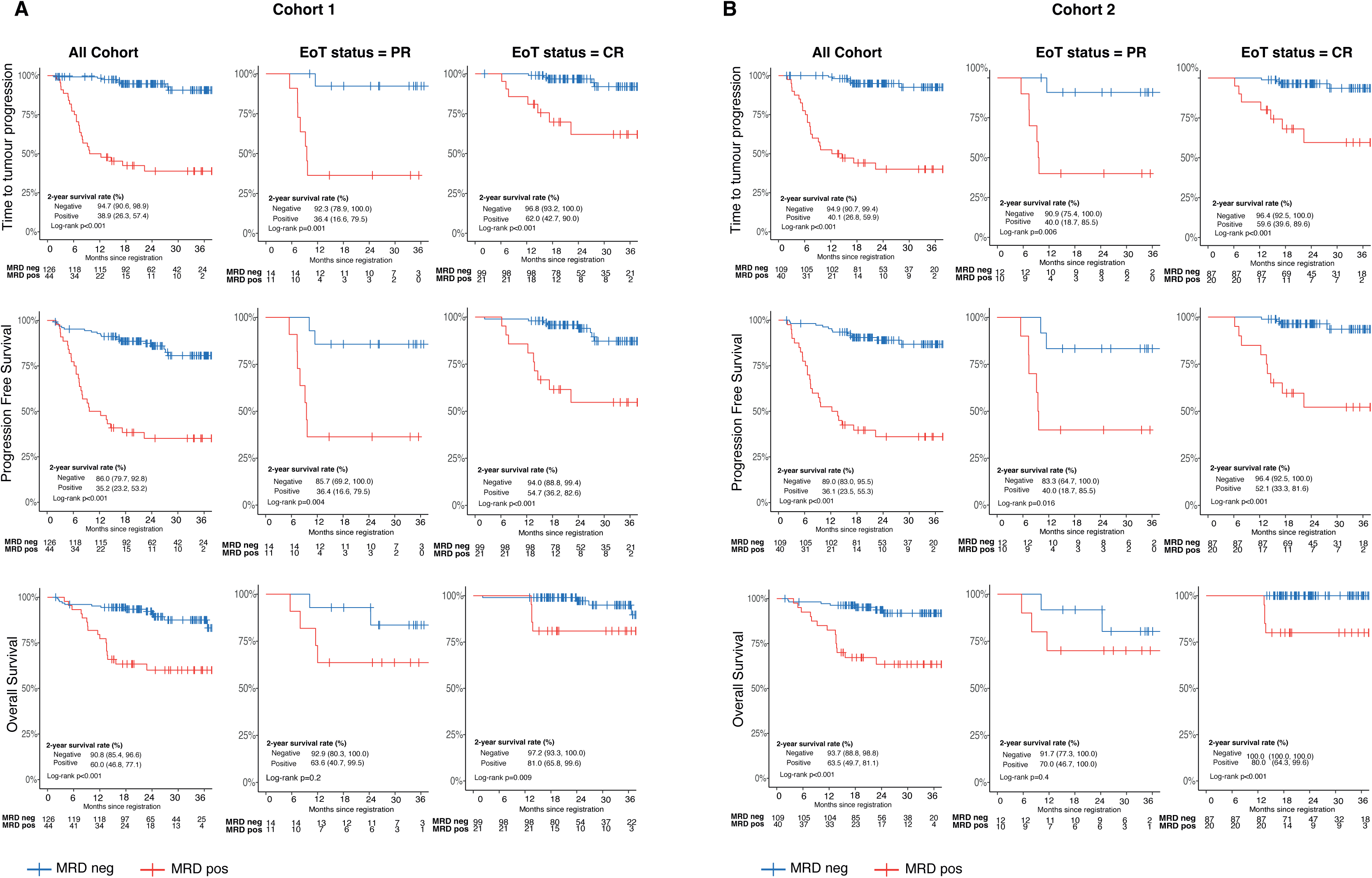
Survival of Cohort 1 and Cohort 2 by EoT MRD status and by EoT radiological response status. A-B. TTP, PFS and OS are shown for the full cohort (lè), then restricted to those with EoT radiological status of PR (center) or CR (right) for Cohort 1 (**A**) or Cohort 2 (**B**). The survival curves are compared with a two-sided log-rank test.

**Supplementary Figure 13.**
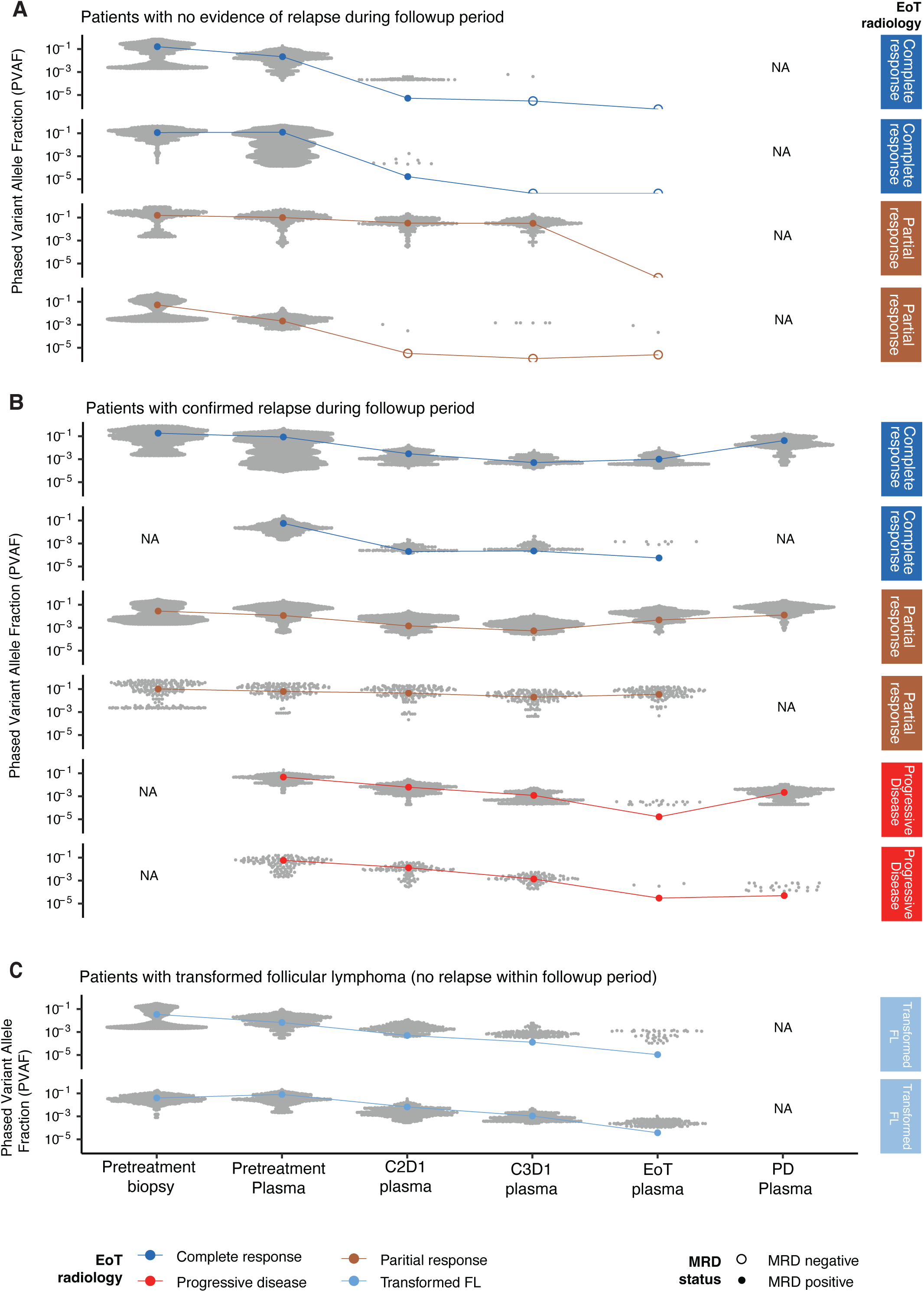
Patterns of response of individual phased variants in selected patient groups. A-C. Examples of PV-supported MRD data in patients with no evidence of relapse during follow-up period (**A**), with confirmed relapse during follow-up period (**B**), or patients with transformed follicular lymphoma with persistent MRD positivity but no evidence of relapse during the follow-up period (**C**). The EoT radiological response status is indicated to the right of each plot. The MRD status is indicated by open (negative) or filled (positive) circles at each 6mepoint.

## Extended Data Tables

Extended Data Table 1 – Design of sequencing panel, gene list and co-ordinates

Extended Data Table 2 – Quality control metrics for all samples included in DIRECT

Extended Data Table 3 – Results of multivariable analysis of pretreatment ctDNA and clinical risk factors

Extended Data Table 4 – List of all somatic variants detected in biopsy samples

Extended Data Table 5 – List of all somatic variants detected in plasma samples

Extended Data Table 6 – Calculator to predict probability of plasma genotyping success

Extended Data Table 7 – List of LymphGen calls in DIRECT cases

Extended Data Table 8 – List of all structural variants identified in DIRECT cases

Extended Data Table 9 – Phased variant numbers, read counts and MRD status by patient

## Notes

### Competing Interest Statement

DJH: reports research funding from Astra Zeneca and GSK.
SB: research funding from Beigene, F Hoffman la Roche and Takeda
SKM: Speaker fees and conference travel: SOBI, Alexion, AZ
DH and VM: employment and stock ownership AstraZeneca

### Author Declarations

The study was reviewed and given favorable opinion by East of England, Cambridge Central Research Ethics Committee (20/EE/0068) and the UK Health Research Authority. All patients provided written informed consent prior to participating. The study was performed in accordance with the spirit and the letter of the declaration of Helsinki, the conditions and principles of ICH E6 Good Clinical Practice, the protocol and applicable local regulatory requirements and laws.

